# Proteome reveals antiviral host response and NETosis during acute COVID-19 in high-risk patients

**DOI:** 10.1101/2022.03.02.22271106

**Authors:** Alina Bauer, Elisabeth Pachl, Johannes C. Hellmuth, Nikolaus Kneidinger, Marion Frankenberger, Hans C. Stubbe, Bernhard Ryffel, Agnese Petrera, Stefanie M. Hauck, Jürgen Behr, Rainer Kaiser, Clemens Scherer, Li Deng, Daniel Teupser, Narges Ahmidi, Maximilian Muenchhoff, Benjamin Schubert, Anne Hilgendorff

## Abstract

SARS-CoV-2 remains an acute threat to human health, endangering hospital capacities worldwide. Many studies have aimed at informing pathophysiologic understanding and identification of disease indicators for risk assessment, monitoring, and therapeutic guidance. While findings start to emerge in the general population, observations in high-risk patients with complex pre-existing conditions are limited.

To this end, we biomedically characterized quantitative proteomics in a hospitalized cohort of COVID-19 patients with mild to severe symptoms suffering from different (co)-morbidities in comparison to both healthy individuals and patients with non-COVID related inflammation. Deep clinical phenotyping enabled the identification of individual disease trajectories in COVID-19 patients. By the use of this specific disease phase assignment, proteome analysis revealed a severity dependent general type-2 centered host response side-by-side with a disease specific antiviral immune reaction in early disease. The identification of phenomena such as neutrophil extracellular trap (NET) formation and a pro-coagulatory response together with the regulation of proteins related to SARS-CoV-2-specific symptoms by unbiased proteome screening both confirms results from targeted approaches and provides novel information for biomarker and therapy development.

**Graphical Abstract:** Sars-CoV-2 remains a challenging threat to our health care system with many pathophysiological mechanisms not fully understood, especially in high-risk patients. Therefore, we characterized a cohort of hospitalized COVID-19 patients with multiple comorbidities by quantitative plasma proteomics and deep clinical phenotyping. The individual patient’s disease progression was determined and the subsequently assigned proteome profiles compared with a healthy and a chronically inflamed control cohort. The identified disease phase and severity specific protein profiles revealed an antiviral immune response together with coagulation activation indicating the formation of NETosis side-by-side with tissue remodeling related to the inflammatory signature.

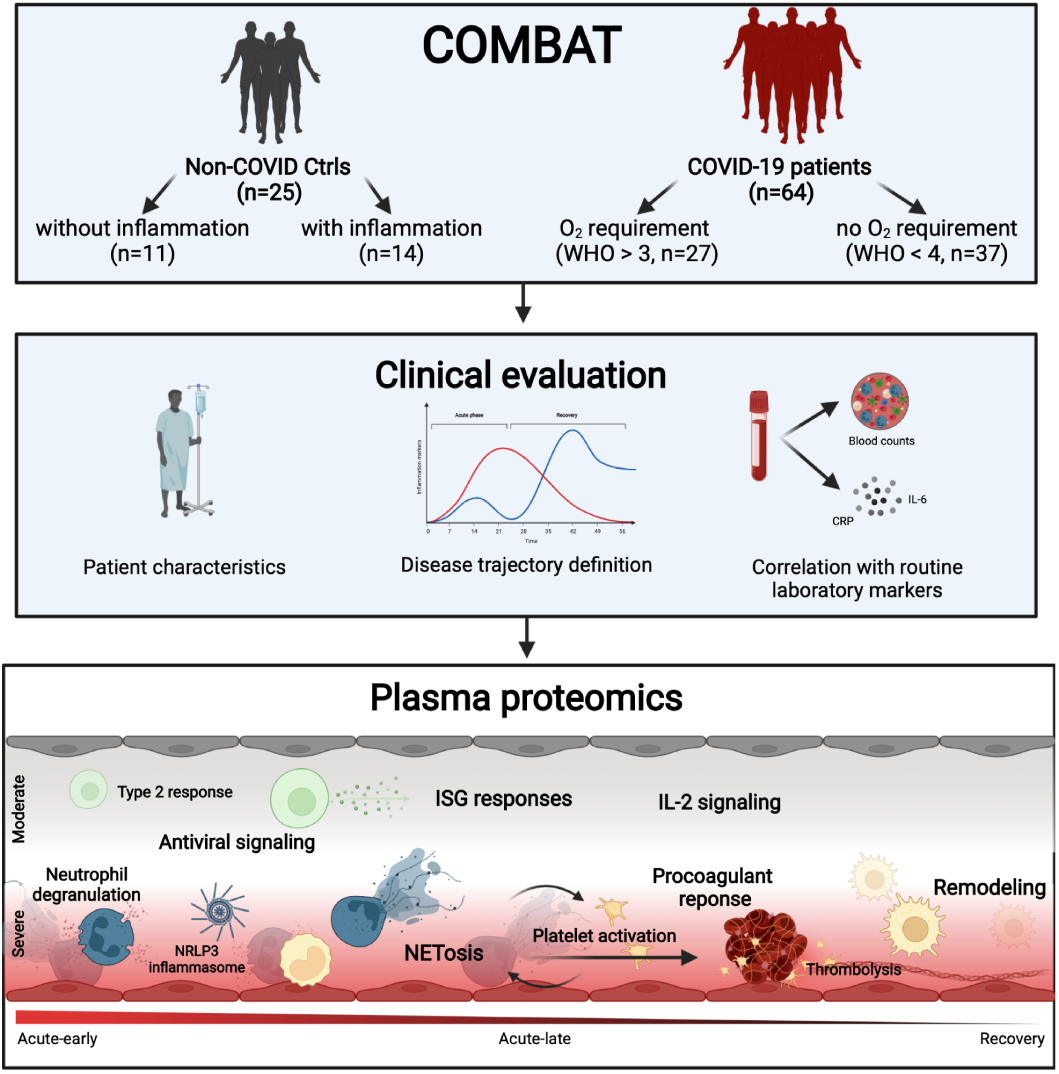

## Introduction

The SARS-CoV-2 pandemic continues to pose an immediate threat to global health. As of January 2022, worldwide COVID-19 cases exceed 250 million and deaths have surpassed 5,4 million (WHO Coronavirus (COVID-19) Dashboard). Clinical manifestations vary from asymptomatic carrier to severe illness, organ dysfunction, chronic health impairment including long-COVID, and death (Williamson *et al*, 2020). To gain deeper insight and inform patient care, epidemiological approaches addressed clinical characteristics of different SARS-CoV-2 infection phases in the overall population and identified risk factors for adverse outcomes such as diabetes or hyperlipidemia (Chen *et al*, 2020b; Huang *et al*, 2020; Guan *et al*, 2020). Studies focused on the identification of clinical signs and early markers that reliably enable monitoring and treatment strategies (Liu *et al*, 2020b; Zhou *et al*, 2020) including nationwide approaches stemming from the United Kingdom, Germany, France, Israel, and the USA (Williamson *et al*, 2020; Nachtigall *et al*, 2020; Piroth *et al*, 2021). While these attempts are already challenging in the general population (Booth *et al*, 2021; Hodges *et al*, 2020; Williamson *et al*, 2020), the aim has yet to be reached in cohorts of high-risk patients characterized by a complex picture of preexisting comorbidities. Care for these patients results in resource-intensive monitoring and treatment and thus remains a critical hurdle even for maximum care hospitals. Poor vaccination response rates in a large number of such complex cases and vaccine breakthroughs further complicate the picture (Boyarsky *et al*, 2021; Malinis *et al*, 2021; Juthani *et al*, 2021).

Characterization of immune phenomena such as the ‘cytokine storm’ (Yang *et al*, 2021; Buszko *et al*, 2021) helped to guide treatment initiation and aided first therapeutic approaches (Chen *et al*, 2020b; Huang *et al*, 2020; Zhang *et al*, 2020a). Changes in human plasma protein levels have been suggested as disease indicators (Shu *et al*, 2020; Messner *et al*, 2020; Park *et al*, 2020), in line with the implementation of protein markers for other viral diseases (Oxford *et al*, 2016). The analyses were furthermore used to gain pathophysiological insight in order to develop new therapeutic strategies (Demichev *et al*, 2021; Haljasmägi *et al*, 2020; Filbin *et al*, 2021).

To increase pathophysiologic insight and enable the identification of disease indicators in high-risk COVID-19 patients with significant preexisting conditions to inform monitoring and treatment decisions in the most important disease phases, we profiled host responses to SARS-CoV-2 infection by the use of quantitative plasma proteomics. Tracing disease trajectories by individual expression of inflammation markers enabled us to improve general time-of-infection-based approaches (Schulte-Schrepping *et al*, 2020).

We thereby successfully identified disease grade and disease phase-specific proteome profiles side-by-side with the regulation of characteristic routine laboratory variables in a high-risk, multimorbid patient cohort. Our study included survivors and non-survivors from COVID-19 and a range from mild to severe disease symptoms compared to patients with acute non-COVID-19 related inflammation, as well as non-inflammatory control cases. We thereby delineated both COVID-19-specific and general immune responses together with the phase-specific involvement of coagulation and remodeling processes as well as the differential regulation of proteins related to SARS-CoV-2-specific symptoms with the potential to significantly inform monitoring and treatment approaches.

## Results

### Assignment to disease severity revealed differences in patient characteristics at admission

The study prospectively enrolled 64 patients with PCR confirmed SARS-CoV-2 infection during the first phase of the COVID-19 pandemic in Germany (03/2020 to 08/2020), before steroid treatment for SARS-CoV-2 was routinely prescribed. Patients were enrolled shortly before or at the onset of the acute infection phase when laboratory signs of infection and disease-specific symptoms develop. Twenty-five patients with acute (inflammatory control group; *Ctrl-infl*) or no/low non-COVID-19 related inflammation (healthy control group; *Ctrl-noninfl*) were additionally included in the study as control groups (see **Materials and Methods - Clinical Data Collection, patient grouping, and disease phase assignment,** and **Table 1**).

**Table 1:** Group and phase definitions as well as study overview.

| Study groups | Description |
| --- | --- |
| COVID-19 (C19; n=64) | PCR positive SARS-CoV-2 infection |
| C19 oxygen group (C19-ox; n=27) | PCR positive SARS-CoV-2 infection; oxygen supply due to COVID-19: WHO score 4-8 |
| C19 no-oxygen group (C19-nonox; n=37) | PCR positive SARS-CoV-2 infection; no oxygen supply due to COVID-19: WHO score 1-3 |
| inflammatory control group (Ctrl-infl; n=14) | no SARS-CoV-2 infection (PCR); patient with diagnosed infection/inflammation CRP >0.5 mg/dl or IL-6 >5.9 pg/ml |
| healthy control group (Ctrl-noninfl; n=11) | no SARS-CoV-2 infection (PCR); CRP ≤0.5 mg/dl and IL-6 ≤5.9 pg/ml |
| **Inflammation peak** | Time of the highest measured CRP or IL-6 value (whichever occurred later) broadened +24h |
| **Disease phase** |  |
| acute-early | Interval between disease onset (symptoms and/or positive PCR test) until 24h post inflammation peak |
| acute-late | Interval starting 24h post inflammation peak until hospital discharge |
| recovery | Time after hospital discharge |
| **Biospecimen** | Heparin plasma |
| **Sample analysis** | Proteomics: Olink Explore 1536/384 |
| **Routine biochemical indices** | Laboratory values: Monocyte count [$\times 10^9/L$], Monocytes [%], Neutrophil count [$\times 10^9/L$], Neutrophils [%], Lymphocyte count [$\times 10^9/L$], Lymphocytes [%], Leukocyte count [$\times 10^9/L$], IL-6 [pg/mL], C-reactive protein (CRP) [mg/dl], Creatinine [mg/dl], Platelets [$10^3/\mu l$], Prothrombin Time, INR [sec], Fibrinogen [mg/dl], high sensitivity TroponinT (hsTroponinT) [$\mu g/l$], Partial thromboplastin time (PTT) [sec], Ferritin [$\mu g/l$], D-Dimer [$\mu g/ml$], Procalcitonin (PCT) [$\mu g/l$], eGFR [ml/min/1.73m<sup>2</sup>] |
| **Comorbidity (groups)** | Diabetes, high cholesterol, cardiovascular disease, lung disease, kidney disease, immuno-compromised status, steroid intake during or before proteomics sampling, superinfection during proteomics sampling |

A maximum WHO score ≥4 (Blueprint, 2020) during the hospital stay was observed in 27 patients (*C19-ox* group) and ≤3 in 37 patients (*C19-nonox* group) (**Figure 1A**). The disease severity groups showed differences in age and gender (age: *C19-ox*: 70 (IQR 59-79); *C19-nonox*: 57 (IQR 48-70); females: *C19-ox*: 37.0%; *C19-nonox* group: 32.4%), whereas no difference was observed for the time between symptom onset and hospitalization (*C19-ox*: 5 (IQR 1-8); *C19-nonox*: 5 (IQR 2-9) (days)) in contrast to a greater length of hospital stay in *C19-ox* patients (*C19-ox*: 12 (IQR 12-56); *C19-nonox*: 10 (IQR 7-17)). Most prevalent symptoms for *C19-ox* and *C19-nonox* patients were dyspnea (59.3%; 37.8%), fever (51.9%; 54.1%), fatigue (51.9%; 29.7%), and dry cough (48.2%; 48.7%). Both *C19-ox* and *-nonox* patients presented with different comorbidities including cardiovascular disease (66.7%; 64.9%), pre-existing lung disease (33.3%; 13.5%), immune compromise (37.0%; 24.3%), diabetes (33.3%; 18.9%), and hyperlipidemia (22.2%; 18.9%). In the course of the disease, some patients suffered from acute kidney failure (*C19-ox*: 11.1%; *C19-nonox*: 2.7%), whereas secondary bacterial, fungal, and/or viral infections (‘superinfection’) occurred more frequently in *C19-ox* patients (*C19-ox*: 44.4%; *C19-nonox*: 27.0%). 22.2% and 44.4% of patients in the *C19-ox* group underwent non-invasive or invasive ventilation. Therapeutic interventions including antibiotic (*C19-ox*: 81.5%; *C19-nonox*: 64.9%) and antithrombotic therapy (*C19-ox*: 59.3%; *C19-nonox*: 62.2%), parenteral nutrition (*C19-ox*: 37.0%; *C19-nonox*: 50.0%), non-opioid analgesics (*C19-ox*: 33.3%; *C19-nonox*: 40.5%) were administered in the majority of C19 patients irrespective of disease severity. Asthma therapy (*C19-ox*: 55.6%; *C19-nonox*: 8.1%) and antiviral treatment (*C19-ox*: 44.4%; *C19-nonox*: 16.2%) were used more frequently in higher disease grades.

**Figure 1.**
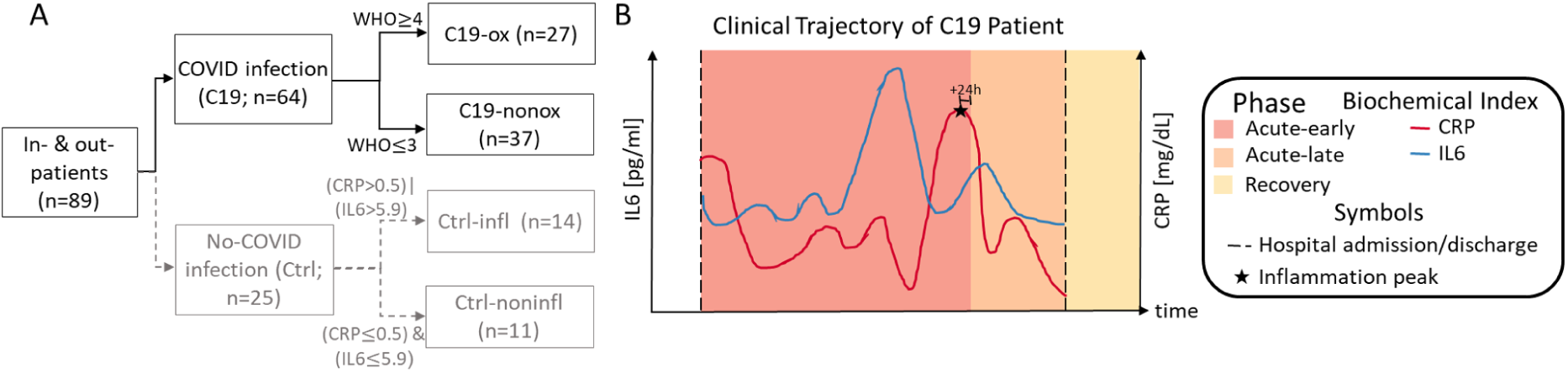
**A:** Patient number per study group considering C19 (black, solid) and non-C19 patients (gray, dashed); **B:** Exemplary COVID-19 disease trajectory based on routine biochemical indices IL-6 (blue) and CRP (red) during the hospital stay (dashed black lines) considering the respective disease phases (*acute-early*, *acute-late,* and *recovery phase*).

*Non-C19* patients were assigned to the two control groups based on levels of the inflammatory markers IL-6 and CRP, *Ctrl-infl* (n=14) and *Ctrl-noninfl* (n=11) (**Table 1**). Median age in years did not differ between both control groups (*Ctrl-infl*: 74 (IQR 61-67); *Ctrl-noninfl*: 69 (IQR 51-75)), whereas female patients were more frequent in the *Ctrl-noninfl* group (*Ctrl-infl*: 14.3%; *Ctrl-noninfl*: 36.7%). The median length of hospital stay (in days) was 9 (IQR 7-12) in *Ctrl-infl* and 1 (IQR 0-2) in *Ctrl-noninfl*. *Ctrl-infl* and *Ctrl-noninfl* patients were characterized by a high prevalence of comorbidities including cardiovascular disease (85.7%; 63.6%), pre-existing lung disease (14.3%; 36.4%), immune deficiency (50.5%; 9.1%), hyperlipidemia (28.6%; 18.2%), and diabetes (21.4%; 18.2%). Patient characteristics at hospital admission for all study groups are presented in **Table S1 & S2**, details about the results of performed statistical tests are shown in **Table S3**.

### Identification of distinct clinical trajectories for disease-severity and -phase assignment

Next to disease-severity assignment according to the WHO criteria (Blueprint, 2020), we grouped samples of the 64 C19 patients in three distinct disease phases - *acute-early*, *acute-late*, and *recovery -* based on the individual trajectory of the inflammation markers IL-6 [pg/ml] and CRP [mg/dl] (**Figure 1B**, **Figure S1**, see **Materials and Methods - Clinical Data Collection, patient grouping, and disease phase assignment**). Forty-four samples of 35 patients were assigned to the *C19 acute-early phase* (interval from disease onset/or first positive PCR test until inflammation peak). Forty-four samples of 40 patients were assigned to the *acute-late phase* (interval from inflammation peak to discharge), whereas 15 samples of 13 patients were assigned to the *recovery phase* (after discharge). For significance tests, one sample per individual was used to avoid autocorrelation (see **Material and Methods - Data analysis**).

This disease phase assignment was found to increase the fit of clinical symptoms and critical events, e.g., ICU admission or the initiation of oxygen treatment to the early disease phase (**Figure 2A** (left)) in comparison to general time-of-infection based approaches that define disease phases by days from symptom onset (0-10 days, early phase; 10 days-discharge, late phase) (Schulte-Schrepping *et al*, 2020) (**Figure 2A** (right)). In addition, we showed CRP and IL-6 values to be statistically different when comparing the *acute-early* and *acute-late* disease phase (CRP: P-value<0.01; IL-6: P-value<0.01), as expected per design of phase assignment, whereas the disease phases based on the general time-of-infection approaches did not show significant differences for these routine markers (CRP: P-value=0.57; IL-6: P-value=0.40), indicating poor separation of subgroups (**Figure 2B**).

**Figure 2:**
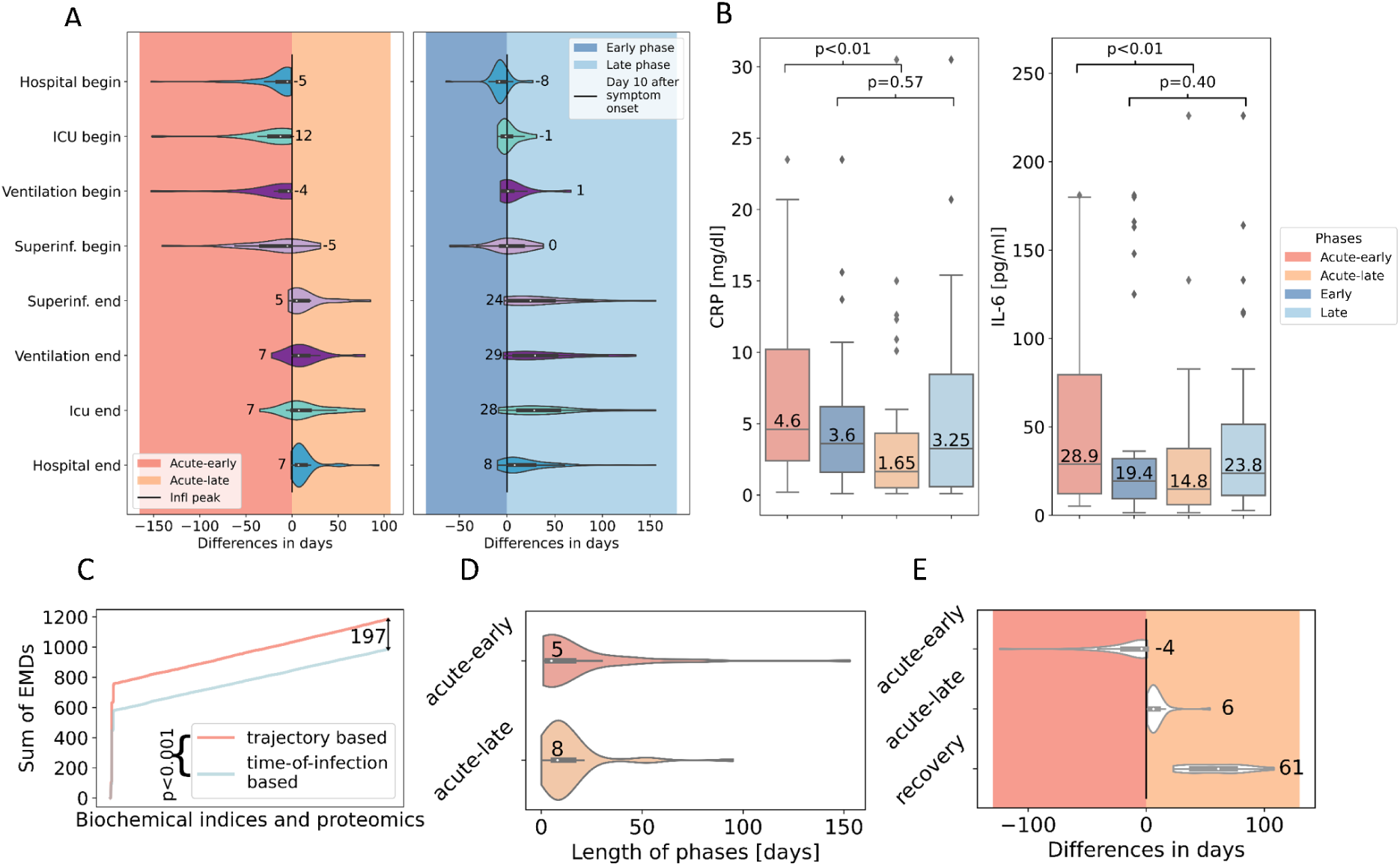
**A:** Difference in days (median) between critical clinical events and disease phase assignment based on the individual trajectory-based approach (left) and the general time-of-infection-based approach (right). ICU: intensive care unit. **B:** Distribution of CRP and IL-6 values in different disease phases based on the individual trajectory-based (red) and general time-of-infection-based (blue) approach for disease phase assignment. **C:** Sum of earth mover’s distance (EMD) from routine biochemical indices and proteomic levels comparing the trajectory-based (red) and time-of-infection-based (blue) approach for disease phase assignment. **D:** Length of disease phases (days; median) on patient level. **E:** Difference in days (median) between sampling time point and inflammation peak.

Quantitatively comparing the discriminatory power of the two approaches, we analyzed the overall earth mover’s distance (EMD) (Rubner *et al*, 2000) between the different phases defined by either the time-of-infection or inflammatory marker-based approach and found a significantly higher EMD (two-sided Wilcoxon Rank-sum test, P-value<0.001) for the inflammatory marker approach, indicating a clearer distinction between the different disease phases (**Figure 2C**).

The median length of the *acute-early phase* was five (IQR 3-16) and the *acute-late* phase was eight (IQR 6-16) days (**Figure 2D**). Sample acquisition for proteomic analysis was performed in accordance with routine procedures between four (IQR 0 - 19) days before to six (IQR 3-10) days after the inflammation peak as determined by Il-6 and CRP expression levels. Post-discharge (*recovery*) samples for proteomic analysis were obtained 61 (IQR 38-75) days after the patient surpassed the inflammation peak (**Figure 2D** **& 2E**). *C19* patients from all groups entered the hospital five (IQR 3-16) days before and were discharged seven (IQR 5-15) days after the inflammation peak. All patients in need of intensive care during their hospitalization were admitted to the ICU 12 (IQR 3-25) days before and discharged seven (IQR 0-20) days after the inflammation peak. In the majority of C19 patients, mechanical ventilation was initiated four days before the inflammation peak (IQR 2-10) and was terminated in the *acute-late phase* eight (IQR 4-21) days after the inflammation peak. Twenty-two *C19* patients developed a secondary (super)infection (**Figure 2A** (left)).

### Disease-severity and -phase-dependent characteristics of routine laboratory values at admission and in the course of disease

At the time of admission, *C19* patients showed significant changes in routine biochemical indices: Patients assigned to severe disease (*C19-ox*) showed comparable elevation of neutrophils when compared to the *Ctrl-infl* group, whereas patients with lower disease grades (*C19-nonox*) were characterized by blood cell counts within the physiologic range apart from monocytosis (**Table S1 & S2**). Whereas CRP levels were found to be significantly different in all other group comparisons, CRP levels were comparable in patients from the *C19-ox* and *Ctrl-infl* group together with elevated levels of procalcitonin (PCT) and fibrinogen, thereby indicating the pathologic but non-discriminatory elevation of these parameters in *C19-ox* patients when compared to patients suffering from inflammation of different origin (**Table S1 & S2**). Fibrinogen levels in *C19-ox* patients showed log2(FC)=0.42 higher abundance when compared to the *C19-nonox* patients. However, ferritin levels were pathologically log2(FC)=1.7 (*C19-ox*) to log2(FC)=2.5 (*C19-nonox*) increased in both *C19* severity groups when compared to *Ctrl-infl* in contrast to lower partial thromboplastin time (PTT, sec) in both C19 disease grades with most pronounced changes in *C19-ox* (log2(FC)=-0.27, **Table S1 & S2**). Reference values for routine laboratory indices are given in **Table S4**.

### Disease severity dependent differences in routine biochemical indices

When investigating the course of the disease, blood cell counts in *C19* patients in the *acute-early* phase of disease were characterized by decreased lymphocyte counts and proportion ([x10^9^/L]: log2(FC)=-0.69, [%]: log2(FC)=-0.54) that increased in the *acute-late* and *recovery* phase, mainly driven by their downregulation in more diseased patients ([x10^9^/L]: log2(FC)=-0.51; [%]: log2(FC)=-1.0) when compared with *Ctrl-infl* (**Figure 3A&C, Figure S2, Table S4**). Whereas *C19-ox* patients in their *acute-early* and *acute-late phase* were comparable to the *Ctrl-infl* group with respect to elevated monocyte proportions, *C19-nonox* patients showed lower levels in the *acute-early* phase (log2(FC)=-3.05) that increase in the *acute-late* phase (log2(FC)=0.58). Likewise, neutrophil proportions were significantly elevated in *C19* patients in the *acute-early* phase compared to *acute-late* phase (log2(FC)=0.13), with a trend to higher neutrophil levels in more diseased patients in the acute disease phase (*acute-early:* log2(FC)=0.12; *acute-late:* log2(FC) = 0.14).

**Figure 3.**
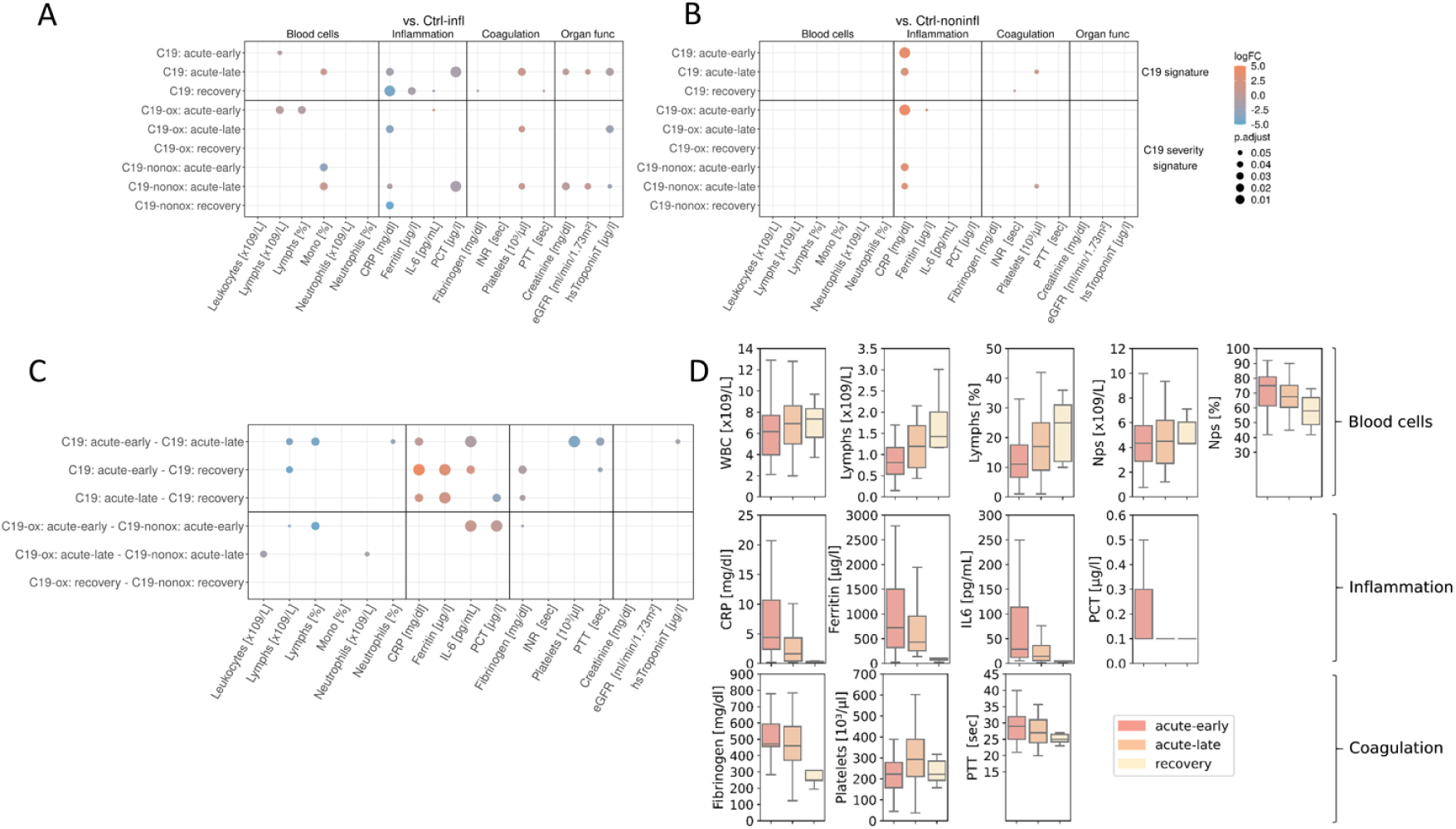
Dot plot of routine biochemical indices revealed significant (P-value ≤ 0.05) differences, describing a general *C19* and a severity-based *C19* signature when compared to the *Ctrl-infl* (**A**) and *Ctrl-noninfl* group (**B**) as well as the trajectory (**C**). Points represent significant level differences in the respective group comparison, where the color of the dots describes the effect size (log2 fold change) and the size of the dots the significance niveau (adjusted P-value). **D:** Distribution of selected biochemical indices over the course of the disease.

In the *acute-late* (*C19-ox*: log2(FC)=-3.54, *C19-nonox*: log2(FC)=-1.16) and *recovery phase* (*C19-ox*: log2(FC)=-4.27, *C19-nonox*: log2(FC)=-4.86) CRP levels were lower in both *C19* severity groups when compared to the *Ctrl-infl* group (**Figure 3**). The overall decline in CRP values in the course of the disease in *C19* patients is most pronounced in more diseased individuals to a level of the *Ctr-noninfl* cohort, whereas in *C19-nonox* patients moderately elevated CRP levels remain in the acute disease phase and normalize in the *recovery* phase together with the *C19-ox* CRP levels (**Figure 3D, Table S5&6**). Likewise, IL-6 values were differentially regulated through the course of the disease (**Figure 3D, Table S5&6**) with *C19-ox* patients in their *acute-early phase* showing significantly higher IL-6 levels (log2(FC)=1.83, **Figure 3C**) when compared to *C19-nonox* patients, in line with recent data (Herold *et al*, 2020).

Likewise, other inflammation parameters normalized in *C19* patients in a disease-severity characteristic manner: While *C19-ox* patients in their *acute-early* phase showed elevated PCT levels that were indistinguishable from the *Ctrl-infl* group whereas *C19-nonox* patients did not show elevated levels in the *acute-early* phase (log2(FC)=1.59; *C19-ox* higher). Patients from both severity groups demonstrated normalized levels as compared to *Ctrl-noninfl*. Additionally, we found a slow normalization of ferritin levels over the course of the disease in all *C19* patients (*C19*: *acute-early* vs. *recovery*: log2(FC)=3.03; C19: *acute-late* vs. *recovery*: log2(FC)=2.30) (**Figure 3C & D**), although ferritin levels still exceeded *Ctrl-infl* levels up until the *acute-late* phase (*acute-early*: log2(FC)=1.42, *acute-late*: log2(FC)=0.76) (**Figure 3B, Table S5&6**).

Accompanying the inflammatory response, platelet counts showed a physiologic niveau in more severely diseased patients in the *acute-early* phase to elevated levels exceeding *Ctrl-infl* levels in the *acute-late phase* (log2(FC)=0.75). In *C19-nonox* patients, the analysis revealed persistently high and even further increasing platelet counts in the course of the disease when compared to the *Ctrl-infl and Ctrl-noninfl* groups (*Ctrl-infl* - *acute-early:* log2(FC)=0.27, *Ctrl-infl* - *acute-late:* log2(FC)=0.43). Elevated fibrinogen levels in the *acute-early* and *-late* phase were observed in both C19 groups compared to *Ctrl-noninfl*, declining in more diseased patients in the *acute-late* phase with *C19-nonox* patients reaching and *C19-ox* patients reaching close to physiologic levels in the recovery phase (**Figure 3C & D, Figure S2**, **Table S5**). Likewise, elevated PTT levels in *C19-ox* patients were comparable to *Ctrl-infl* in *acute-early* and normalized to physiological levels in the *acute-late* phase. In *C19-nonox* patients, moderately higher PTT levels compared to *Ctrl-noninfl* in *acute-early* normalized during the *acute-late* phase (**Figure 3, Table S5&6**).

Levels for kidney function, *i.e.,* creatinine levels, and estimated glomerular filtration rate (eGFR) or cardiac injury, *i.e.*, hsTroponinT did only show significant differences in the *acute-late* phase when compared to *Ctrl-infl* with creatinine and hsTroponinT reduced (creatinine: log2(FC)=-0.22, hsTroponinT: log2(FC)=-1.32) and eGFR elevated (log2(FC)=0.29) mainly driven by changes in *C19-nonox* patients (**Figure 3A & C, Figure S2**, **Table S5 & 6**).

### Proteomic profiling tracking disease indicators for disease stage and severity

By comprehensively dissecting systemic immune responses at the protein level in C19 patients we significantly added to pathophysiologic insight and enabled the identification of future disease indicators using plasma proteomics obtained on the Olink® Explore platform (see **Materials and Methods - Olink plasma proteomics**). Disease characterization by these means is outlined in the following chapters.

### Type 2 and antiviral immune response in early disease *a*ssociated with subsequent activation of the coagulation cascade in C19 patients

The comparison to *Ctrl-noninfl* patients identified the differential regulation of 356 (*acute-early*) and 261 (*acute-late*) proteins in *C19* patients irrespective of disease severity. The majority of proteins were found to be upregulated (*acute-early*: 341; *acute-late*: 247) and showed a strong overlap of 195 differentially abundant proteins between the disease phases (**Figure 4A & B**, **Figure 5, Figure S3, Table S7**) together with higher absolute log-fold changes in the *acute-early* comparison (*acute-early*: 1.0727 [iQR 0.833 - 1.362]; *acute-late*: 0.9683 [IQR: 0.769- 2.294]). In contrast, the comparison of the *recovery phase* with *Ctrl-noninfl* did not reveal significant differences, indicating a normalization of the pathophysiologic changes after discharge in all patients.

**Figure 4:**
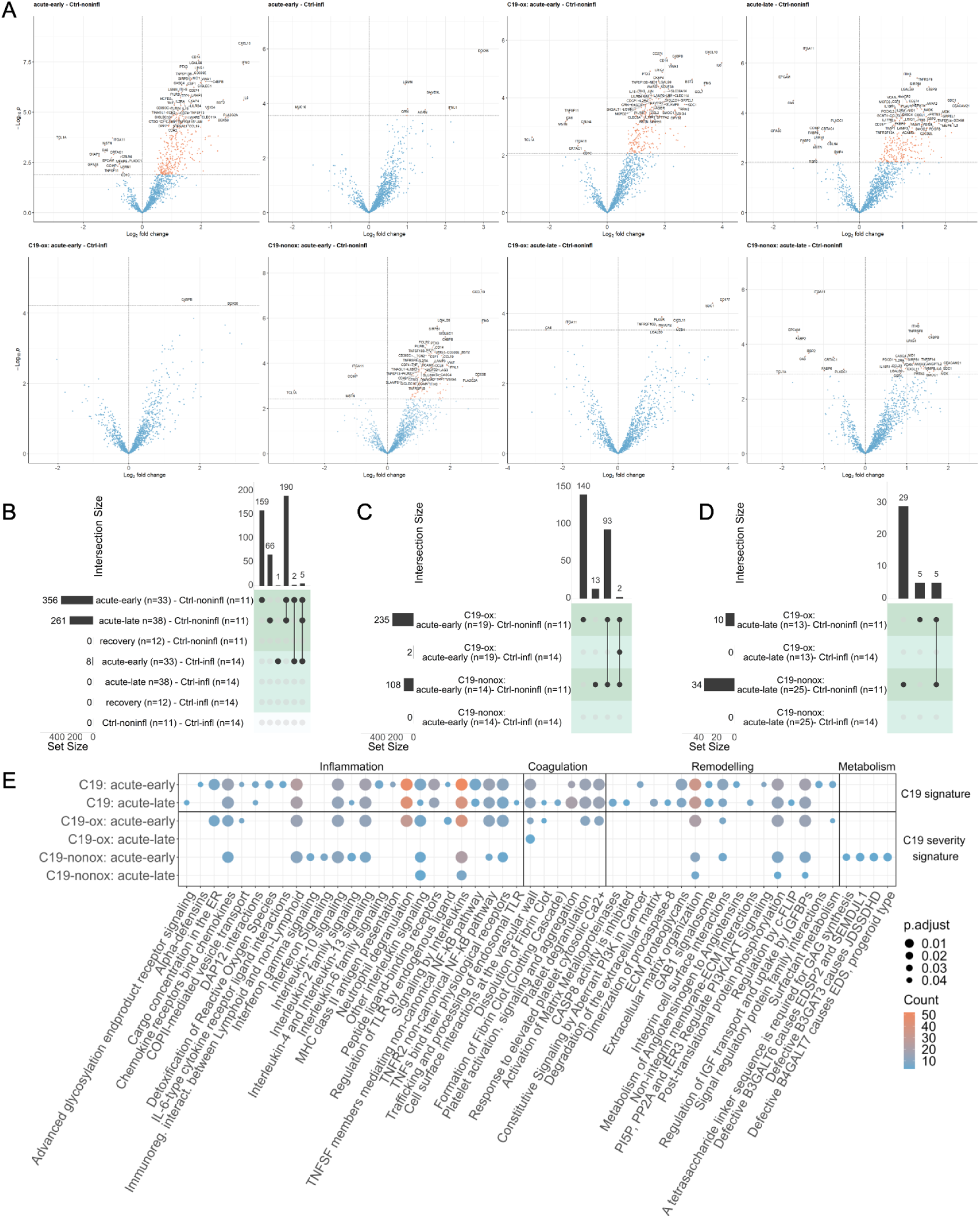
**A:** Volcano plot of all phase comparisons with *Ctrl-noninfl* and *Ctrl-infl* in both severity grades *C19-ox* and *C19-nonox.* **B:** Overlapping significantly differentially abundant proteins in all phase comparisons with *Ctrl-noninfl* and *Ctrl-infl* overall; **C**: between *C19-ox* and *C19-nonox* in *acute-early* phase; **D**: between *C19-ox* and *C19-nonox* in *acute-late* phase. Horizontal bar graphs indicate total differentially abundant proteins (after multiple testing corrections), while vertical bar graphs indicate the number of overlapping differential proteins between different comparisons. **E**: Overlap of enriched pathways between all phase comparisons with *Ctrl-noninfl*.

**Figure 5:**
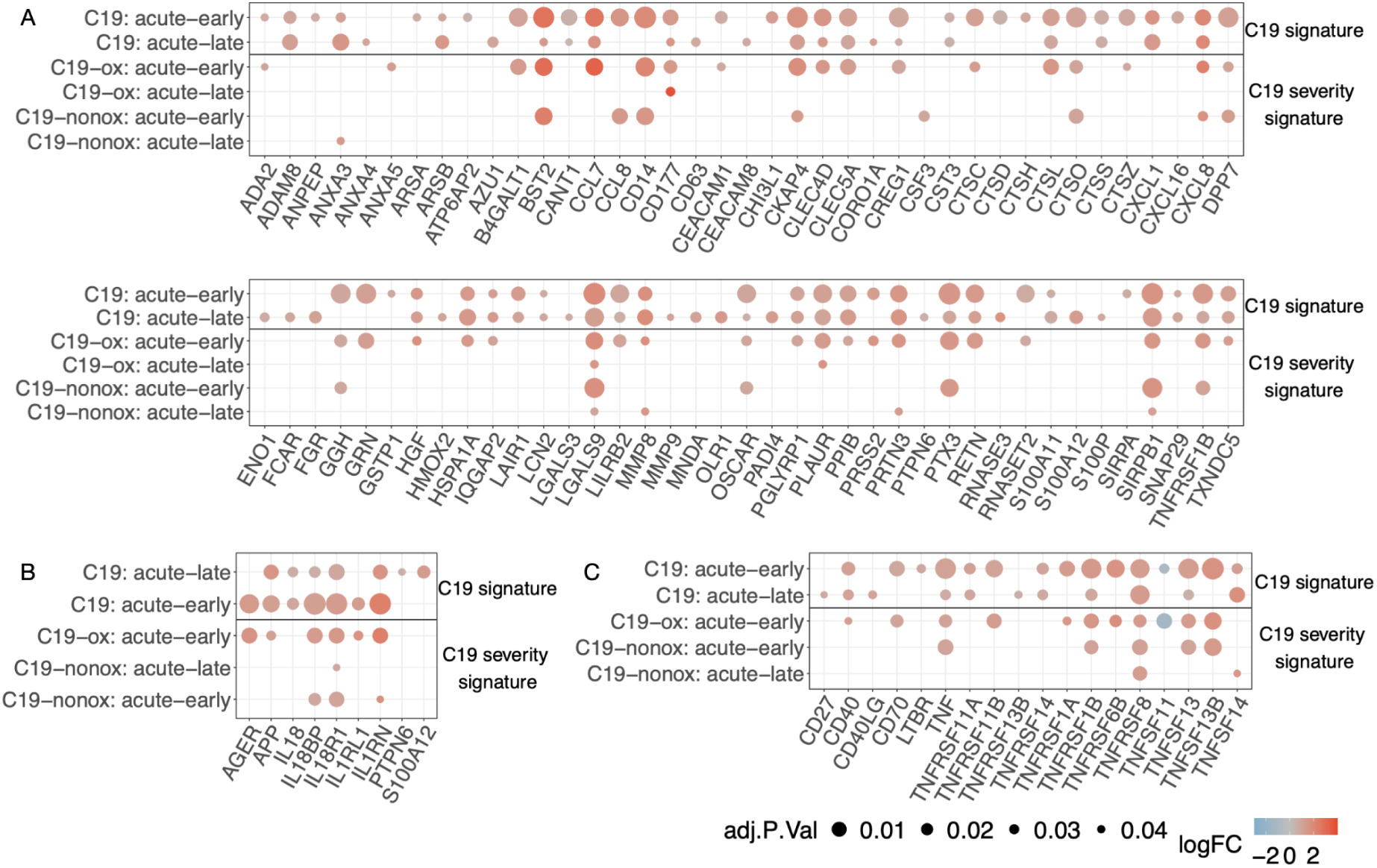
Proteins involved in neutrophil degranulation and (**A**) NET formation, (**B**) IL-1 signaling, (**C**) TNFR cascade in all phase comparisons with *Ctrl-noninfl*.

Protein regulation in both *acute* phases was characterized by strong activation of innate and adaptive immune responses including a type-2 immune response, *i.e., Interleukin-10 signaling*, *Interleukin-4*, and *Interleukin-13 signaling,* and TNFR2 non-canonical NF-kB pathway signaling (*i.e.*, *TNFs bind their physiological receptors*; *TNF receptor superfamily (TNFSF) members mediating non-canonical NF-kB pathway*), *Neutrophil degranulation*, *DAP12 interactions*, *Immunoregulatory interactions between a lymphoid and a non-lymphoid cell* and GPCR signaling (*i.e.*, *Chemokine receptors bind chemokines*; *Peptide ligand-binding receptors*) when compared to *Ctrl-noninfl* (**Figure 4E, Table S8**). The inflammation markers LGALS9 *(Signaling by Interleukins*, *Interleukin-2 family signaling*) and SIRPB1 (*Neutrophil degranulation*, *Signal regulatory protein family interactions*, *DAP12 interactions*) were significantly upregulated in both *acute phases* compared to *Ctrl-noninfl*, next to LRIG1, not represented in any regulated pathway. Interestingly, regulated proteins such as LGALS9 or SIRPB1 hold matrix remodeling capacities next to immune functions (Hsu *et al*, 2020; Chen *et al*, 2019b).

Protein regulation in the *acute-early* phase was uniquely characterized by the involvement of inflammatory processes including interleukin signaling (*i.e.*, *Interleukin-6 family signaling*; *IL-6-type cytokine receptor-ligand interactions*), innate (*i.e.*, *Alpha-defensins*; *Regulation of TLR by endogenous ligand*), and adaptive immune responses (*i.e.*, *MHC class II antigen presentation*), while *Interleukin-2 family signaling* and innate immune response pathways such as *Advanced glycosylation endproduct receptor signaling* and *Trafficking and processing of endosomal TLR* were only significantly regulated in the *acute-late phase*. The differential regulation between the *acute-early* and *-late* phase additionally comprised stress response mechanisms indicated by *Detoxification of Reactive Oxygen Species*, *COPII-mediated vesicle transport,* and *Cargo concentration in the ER* significantly regulated in the *acute-early* disease phase. The *acute-early* phase was furthermore characterized by regulation of the top-ranked inflammation marker CXCL10 (log2(FC)>2, *Signaling by Interleukins*, *Interleukin-10 signaling*, *Chemokine receptors bind chemokines* and *Peptide ligand-binding receptors*), PTX3, involved in *Neutrophil degranulation* (Daigo *et al*, 2016), and CD300E (*Immunoregulatory interactions between a Lymphoid and a non-Lymphoid cell*, *DAP12 interactions*), all persisting but less pronounced in the *acute-late phase* irrespective of disease grade. Likewise, IFNG (log2(FC)>2, *Signaling by Interleukins*), TNFSF13B (*TNFs bind their physiological receptors*, *TNFR2 non-canonical NF-kB pathway*, *TNF receptor superfamily (TNFSF) members mediating non-canonical NF-kB pathway)*, and CD14 (*Regulation of TLR by endogenous ligand*) were predominantly regulated in the *acute-early* phase in all *C19* patients, whereas no differential regulation was observed in the *acute-late* phase when compared to *Ctrl-noninfl*. While TNFRSF8 *(TNFs bind their physiological receptors*, *TNFR2 non-canonical NF-kB pathway*) and HAVCR2 (*Signaling by Interleukins*, *Interleukin-2 family signaling*) were predominantly upregulated in the *acute-late* phase, their regulation was less pronounced in the *acute-early* phase when compared to *Ctrl-noninfl*.

Regulation of coagulation processes could be observed during both acute disease phases when compared to Ctrl-noninfl (i.e., Cell surface interactions at the vascular wall, Platelet activation, signaling and aggregation; Response to elevated platelet cytosolic Ca2+ and Platelet degranulation), accompanied by the upregulation of the coagulation marker C4BPB, not represented in any pathway, whereas Formation of Fibrin Clot (Clotting Cascade) and Dissolution of Fibrin Clot were significantly regulated in the acute-late phase, together with a downregulation of EPCAM (Cell surface interactions at the vascular wall) and upregulation of ITIH3 (Platelet degranulation, Response to elevated platelet cytosolic Ca2+, Platelet activation, signaling and aggregation), present but less pronounced in the acute-early phase, in line with observations that thrombotic events tend to appear in later disease stages of COVID-19 (Al-Ani et al, 2020; Bussani et al, 2020).

Interestingly, a large proportion of proteins associated with neutrophil activity and NET formation such as ANXA3, CCL7, HSPA1A, LCN2, LGALS9, MMP8, PPIB, PRTN3, and RETN (Wang *et al*, 2020; Brinkmann *et al*, 2004) were upregulated in both *acute* phases (*acute-early*: 58/78, *acute-late*: 54/78) compared to *Ctrl-noninfl* with 36 proteins similarly regulated across both *acute* disease phases, confirming previous reports of increased NETosis in *C19* patients by unbiased proteome screening (Bardoel *et al*, 2014; Middleton *et al*, 2020; Nicolai *et al*, 2020; Zhou *et al*, 2021; Smet *et al*, 2021) (**Figure 5A**). Similarly, the differential regulation of *TNF (acute-early: 15/31, acute-late: 11/31)* and *IL-1 (acute-early: 7/23, acute-late: 7/23) signaling pathway* associated proteins were detected in the *acute* disease phases when compared to *Ctrl-noninfl* (**Figure 5B & C**). The proteins point towards the activation of inflammasome-associated processes (Zheng *et al*, 2020) which play an important role in NET formation (Münzer *et al*, 2019, 2021).

Proteins involved in remodeling and repair processes were differentially expressed in both *acute* disease phases when compared to *Ctrl-noninfl* as indicated by the regulation of *Insulin-like Growth Factor (IGF) transport and uptake by Insulin-like Growth Factor Binding Proteins* (IGFBPs), complemented by proteins involved in the *Extracellular matrix organization*, such as NID1 including regulation of *ECM proteoglycans* and *Integrin cell surface interactions*, eGFR signaling (*i.e.*, *GAB1 signalosome*), and *Post-translational protein phosphorylation.* Whereas remodeling processes in the *acute-early* phase were characterized by tissue-cell (*Metabolism of Angiotensinogen to Angiotensins*; *Surfactant metabolism*, PI3K/AKT signaling via, *i.e.,* PI5P, PP2A, and IER3) and cell-cell communication and remodeling (i.e., *Signal regulatory protein family interactions, i.e.,* SIRPB1/SFTPA2/SFTPA1/SIRPA), the protein signature in the *acute-late* phase was dominated by processes such as the *Degradation of the extracellular matrix*, *Activation of matrix metalloproteinases*, *Non-integrin membrane-ECM interactions*, and regulation of the apoptosis pathways (*i.e.*, *CASP8 activity is inhibited*, *Regulation by c-FLIP*, *Dimerization of procaspase-8*). These processes were accompanied by a decrease in the expression of ITGA11 (*Extracellular matrix organization*, *Integrin cell surface interactions*, top-ranked protein in the *acute-late phase*) and the increased expression of SDC1 (*Signaling by Interleukins*, *Extracellular matrix organization*, *Other interleukin signaling*, *Cell surface interactions at the vascular wall*). Not represented in the enriched pathways, the metabolic marker CA6 was found to be regulated in the *acute-late* phase. The downregulation of CA6 is strongly linked to low salivary zinc concentrations, associated with decreased taste acuity (hypogeusia) (Shatzman & Henkin, 1981), and has been used in the diagnosis of Early Sjögren’s Syndrome (Jin *et al*, 2019).

**In summary,** the proteomic response of *C19* patients during the *acute* disease phase was characterized by the activation of classical inflammatory pathways, combined with markers indicating activation of the coagulation cascade and matrix remodeling when compared to *Ctrl-noninfl* individuals (**Figures 4 & 5**; **Figures S3; Tables S7 & 8**). The unique combination of these processes together with the presence of specific markers, *i.e.*, ANXA3, CCL7, HSPA1A, LCN2, LGALS9, MMP8, PPIB, PRTN3, and RETN pointed towards neutrophil degranulation and neutrophil extracellular trap (NET) formation in the *acute* disease phases in *C19* patients, in line with previous studies describing innate immune cell activation in severe COVID-19 including neutrophil degranulation, NETosis, as well as pro-inflammatory / HLA-DRlo monocyte expansion (Bardoel *et al*, 2014; Middleton *et al*, 2020; Nicolai *et al*, 2020; Zhou *et al*, 2021; Smet *et al*, 2021) (**Figure 5**). In addition, we detected a strong activation of interleukin signaling including activation of TNF signaling and a type-2 inflammation with the potential to counteract TNF-related signaling, especially in monocyte-related functions. These changes occurred together with the activation of both cytotoxic and humoral related immune defense mechanisms related to Interleukin-2 family signaling and DAP-12 in the *acute-late* phase, indicating the development of an adaptive immune response (Spolski *et al*, 2018).

Whereas angiotensinogen, surfactant and SIRP metabolism, ROS regulation, and IL-6 signaling dominated protein regulation in the *acute-early* disease phase, the *acute-later* course was characterized by the differential expression of proteins indicating matrix degradation and apoptosis (**Figure 4E**). Pathway enrichment analysis reflected vascular activation and organ damage that persisted into the *acute-late* phase together with markers of both coagulation and thrombolysis along with platelet degranulation in both *acute-early* and *acute-late* disease (**Figure 4E**). However, protein regulation associated with coagulation processes was more pronounced in the *acute-late* phase.

### Confirmation of the antiviral immune response as a C19-characteristic pattern of protein regulation

The comparison of *C19* patients in the *acute* disease phases with *Ctrl-infl* patients demonstrated the differential regulation of eight proteins, all regulated in the *acute-early* phase irrespective of disease severity, whereas the comparison of the *acute-late* phase demonstrated no differentially expressed proteins (**Figure 4A & B, Figure S3, Table S7**). The *C19*-specific profile in the *acute-early* phase demonstrated the upregulation of antiviral signaling (DDX58, IFNL1, SAMD9L, GRN), lysosomal protein degradation (LGMN, TPP1), and Toll-like receptor signaling (Pišlar *et al*, 2020), as well as epithelial cell injury through upregulation of AGRN and downregulation of MUC16, as previously described in COVID-19 (Smet *et al*, 2021) of which a significant proportion was also regulated in the comparison to the *Ctrl-noninfl* group, *i.e.,* DDX58, SAMD9L, LGMN, TPP1, and AGRN regulated in both *acute* phases; IFNL1 and GRN regulated in the *acute-early* phase. However, no coagulation markers or interleukin signaling-associated proteins were found differentially abundant.

The differential regulation of DDX58 (*RIG-I*) controls the recognition of infected cells while IFNL1 leads to the activation of the JAK/STAT signaling pathway resulting in the expression of IFN-stimulated genes (ISGs). Interestingly, these ISGs mediate the antiviral state essential for containment of SARS-CoV-2 in the upper respiratory tract (Pekayvaz *et al*, 2021), while loss of ISG function is associated with severe COVID-19 (Bastard *et al*, 2020; Zhang *et al*, 2020b). The host response is further characterized by the regulation of apoptosis, cell cycle arrest, and DNA damage through SAMD9L, the activation of defense mechanisms involving monocyte differentiation and *MHC class II presentation* through LGMN, lysosomal protease functions controlled by TPP1 and activated through acidification such as GRN that holds a role in inflammation previously associated with COVID-19. Epithelial cell damage was indicated by the regulation of AGRN, as part of the lung basal membrane and MUC16, controlling mucus secretion and engaged in epithelial cell replication and apoptosis.

**In summary,** specific protein regulation in C19 likely related to an activation of the immune system not reflected by routine laboratory variables (*i.e.*, CRP, IL-6, PCT, ferritin, and neutrophil proportions) that did not distinguish *C19-ox* patients from *Ctrl-infl* patients. A strong antiviral immune response side-by-side with markers indicating apoptosis and DNA damage both confirm previous findings as well as delineates the *C19* immune response in the early course of the disease.

### Protein regulation in C19 patients indicates the regulation of antiviral immune response and a NETosis related activation of the coagulation cascade in more severe disease

When analyzing the general inflammatory response based on the comparison to *Ctrl-noninfl* cases, the *acute-early phase* in *C19-ox* patients revealed 235 significantly regulated proteins, with the majority, *i.e.*, 227 proteins showing a strong upregulation in the *acute-early phase* in *C19-ox* patients (**Figure 4A & C**, **Figure 5,Figure S3, Table S7 & S8**). 140 of these 235 differentially regulated proteins characterized the general inflammatory early response in *C19-ox* patients and were not found to be differentially abundant within the early response in *C19-nonox* patients. Specific processes in higher disease severity grades of the *acute-early* phase involved regulation of the innate immune system via *Endogenous ligand TLR signaling* and *Neutrophil degranulation* (supported by an upregulation of the oxygen-specific markers SFTPA2, LBP, GRN, B4GALT1, and RETN in the *acute-early phase*) as well as stress response regulation indicated by enrichment of the pathways *ER to Golgi Anterograde Transport via COPII-mediated vesicle transport* and *Cargo concentration in the ER*. The top 10 of the 140 regulated proteins furtherincluded CCL7 (log2(FC)>2), CCL2, JUN, AGER, NDUFS6 (log2(FC)>2), indicating the activation of inflammatory and oxidative stress mechanisms in severe disease.

Regulated processes in severely affected C19 patients included both prothrombotic and thrombolytic processes such as Cell surface interactions at the vascular wall, Response to elevated platelet cytosolic Ca2+, Platelet degranulation, and Dissolution of Fibrin Clot. Tissue-cell interaction was represented by Surfactant metabolism, accompanied by the upregulation of the oxygen-specific marker SFTPA2 (also involved in Regulation of TLR by endogenous ligand; top 10) in the acute-early phase. All mentioned pathways apart from Dissolution of Fibrin Clot were found to be regulated in the overall comparison including both severity grades (**Figure 4E, Table S8**), implying a strong contribution to the inflammatory and prothrombotic phenotype by their upregulation in patients with severe disease.

The *acute-late* phase in *C19-ox* patients revealed the differential regulation of ten proteins, eight of which were upregulated (**Figure 4A & D, Figure S4, Table S7 & S8)**. Inflammation markers in the *acute-late phase* of *C19-ox* patients included the upregulation of CXCL11 (log2(FC)>2), HAVCR2, TNFRSF10B, SDC1 (log2(FC)>2), and LGALS9, again pointing to a pronounced immune response as well as the activation of remodeling processes.

The coagulation process *Cell surface interactions at the vascular wall* was found to be persistently regulated in *C19-ox* patients in the *acute-late* phase, accompanied by the upregulation of the coagulation marker PLAUR, significantly involved in NET formation (Stavrou *et al*, 2018). Likewise, involvement of remodeling as well as metabolic processes were indicated by the upregulation of CD177, MZB1, and CA6 (all three with log2(FC)>2), as well as by the downregulation of ITGA11 in these patients. Several proteins, including the activation marker CD177, the thrombolytic PLAUR, as well as proinflammatory factors such as TNFRSF10B, HAVCR2, and MZB1 were uniquely regulated in *C19-ox* patients during the *acute-late phase*, in contrast to *C19-nonox* patients.

The comparison of protein regulation in *C19-ox* with patients from the *Ctrl-infl* group revealed the upregulation of DDX58 (log2(FC)>2) and C4BPB in the *acute-early* phase (**Figure 4A & C, Figure S3**), whereas the analysis in the *acute-late* phase did not reveal differentially regulated proteins.

Protein regulation in patients with less severe disease (*C19-nonox*) revealed 108 differentially expressed proteins in the *acute-early* phase when compared to *Ctrl-noninfl*, with an upregulation in the majority of proteins, *i.e*., 104 (**Figure 4A & C, Figure S3, Table S7 & S8**).

Inflammatory processes in the *acute-early phase of C19-nonox* patients comprised the enrichment of *Interleukin-2 family signaling*, *Interferon-gamma (),* and *Interferon signaling* (both unique in this comparison) when compared to *Ctrl-noninfl*. Inflammatory proteins regulated in *acute-early C19-nonox*, but not in *C19-ox* comprised the increased abundance of proteins associated with *Interleukin-10 signaling* (CCL3, CSF3), *Chemokine receptors bind chemokines* (CCL3, CCL21), and proteins not associated with enriched pathways (CCL8, CLEC6A, VSTM1). Similarly, PDGFRA was only differentially regulated in *C19-nonox* but not in *C19-ox*, whereas coagulation activation was not observed by pathway enrichment analysis.

Remodeling processes were indicated by the unique enrichment of pathways in the *C19-nonox* patients that related to Glycosaminoglycan metabolism (*i.e.*, *A tetrasaccharide linker sequence is required for GAG synthesis*; *Defective B3GAT3 causes JDSSDHD*; *Defective B3GALT6 causes EDSP2 and SEMDJL1*; *Defective B4GALT7 causes EDS, progeroid type*). This was accompanied by the downregulation of the remodeling marker COMP(Extracellular matrix organization) as well as by the upregulation of TYMP, C1QTNF1, SCARB2, SIAE, and WISP2, all regulated in *acute-early C19-nonox*, but not in *C19-ox* when compared to *Ctrl-noninfl*. All other significantly enriched pathways in this comparison were shared with the profile observed in the *acute-early phase* of *C19-ox* patients (**Figure 4E, Table S8**). When compared to *Ctrl-noninfl*, the *acute-late* phase in *C19-nonox* patients revealed 34 significantly regulated proteins, 25 of which were upregulated (**Figure 4D, Figure S3, Table S7 & S8**). Specifically, 29 of those 34 proteins were regulated in the *acute-late* phase of *C19-nonox* patients only, with the top 10 ranked proteins including markers of inflammation (TNFRSF8, LRIG1), coagulation (EPCAM, C4BPB, ITIH3), remodeling (CRTAC1, NID1), and metabolic processes (FABP2, RBP2, CASC4). Pathway enrichment analysis revealed the regulation of inflammatory processes including cytokine signaling via *Other interleukin signaling*. Remodeling processes in *C19-nonox* patients during the *acute-late phase* included *Regulation of Insulin-like Growth Factor (IGF) transport and uptake by Insulin-like Growth Factor Binding Proteins (IGFBPs)*, *Post-translational protein phosphorylation*, and *Extracellular matrix organization*.

All differentially expressed pathways in *C19-nonox* patients that were regulated in the disease independent analysis showed a shared expression with *C19-ox* patients indicating an equal contribution of both disease severities to the independent comparison in the acute-early disease phase. In contrast, the comparison demonstrated a dominating impact of the *C19-nonox* patients on the general signature in the acute late phase.

The protein profile in lower disease severity grades showed no differentially regulated proteins when comparing the *acute-early* and the *acute-late phase* to the *Ctrl-infl* group (**Figure 4D**). Similarly, no differentially regulated proteins could be identified in the *recovery* phase for both oxygen-dependent and independent patients (**Figure 4B**).

**In summary,** processes identified in *C19* patients in general as outlined above were found to be more prominently or solely regulated in severely diseased *C19* patients. When comparing severity groups, these changes were accompanied by a strong induction of an innate immune response in the *early-acute* phase indicated by *Regulation* of *TLR through endogenous ligands*, *Neutrophil degranulation,* and stress response mechanisms with *Cargo concentration in the ER,* and *COPII-mediated vesicle transport.* Simultaneously, procoagulant and thrombolytic phenomena, *i.e.*, *Response to elevated platelet cytosolic calcium*, *Platelet degranulation, Response to elevated platelet cytosolic Ca2+, Cell surface interactions at the vascular wall,* and *Dissolution of Fibrin Clot*, could be observed in *C19-ox* patients (**Figure 4E**).

In contrast, protein regulation in less severely diseased *C19* patients was dominated by a general inflammatory response exemplified by the group-specific regulation of interleukin-2 and IFNG signaling in the *acute-early* phase, as well as a shared pattern, *i.e.,* regulated in both severity groups that included remodeling processes indicated by regulation of *Extracellular matrix organization*, *Post-translational protein phosphorylation,* and *Regulation of Insulin-like Growth Factor transport and uptake by Insulin-like Growth Factor Binding Proteins* in the *acute-early* and -*late* phase (**Figure 4E**). Interestingly, no pathways or proteins directly related to coagulation were found to be significantly regulated in less diseased patients, confirming previous clinical observations in these patients (Nicolai *et al*, 2020).

Changes observed in routine laboratory variables were reflected in protein expression patterns, *i.e.*, elevated neutrophil numbers in more diseased patients correlated with the increased presence of markers for neutrophil degranulation and coagulation.

### Dynamic regulation of immune and remodeling processes in the course of disease depending on disease severity

Next, we investigated the disease phase-dependent regulation of plasma proteins in *C19* patients. Changes over the entire disease trajectory revealed 45 inflammation markers, four coagulation markers, 20 markers indicating remodeling processes, and 19 metabolic markers to be differentially regulated (**Figure 6A**, **Figure S3, Table S9 & S10**).

**Figure 6:**
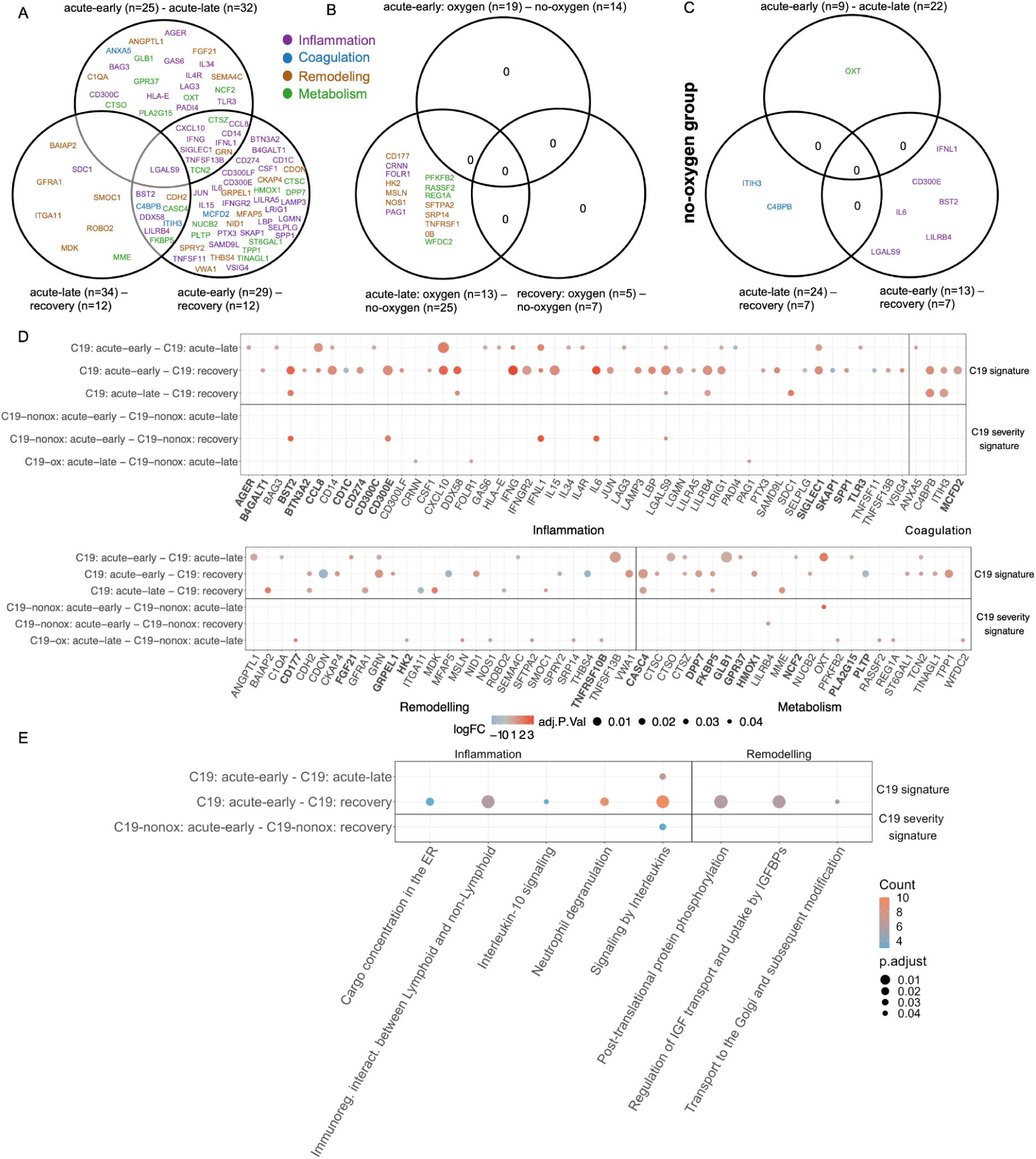
**A:** COVID-19 phase-dependant differential regulation of plasma proteins and their intersections. **B:** Differentially regulated plasma proteins and their intersections for disease severity comparisons (*C19-ox/C19-nonox*) considering different disease phases; purple: inflammation; blue: coagulation; brown: remodeling; green: metabolism **C:** Intersections of differentially abundant plasma proteins for different disease phases in *C19-nonox*. **D**: Log2 fold-change of phase-regulated proteins. Protein symbols in bold are associated with NETosis, IL-1, or TNF signaling. **E:** Overlapping pathways of all disease phase comparisons.

Specifically, 33 proteins were found to be differentially abundant between the *acute-early* and *acute-late* phase, yielding significant differences in interleukin signaling (**Figure 6A** **& B & E & F, Figure S3).** As expected, 17 of those 33 proteins showed strong differences between the *acute-early* and *acute-late* phase when compared to *Ctrl-noninfl*. Here, CD14, IFNG, TNFSF13B, CTSO, ANGPTL1, GRN, C1QA, AGER, IFNL1, LAG3, HLA-E, CCL8, GAS6, IL4R, CTSZ, and PLA2G15 were upregulated in the *acute-early* phase when compared to *Ctrl-noninfl* but were not significantly regulated in later phases, indicating an innate immune host response that involves vascular and matrix remodeling specific proteins and prominent interferon-related signaling in the *acute-early* disease phase. In contrast, PADI4 was only upregulated in the *acute-late* phase when compared to *Ctrl-noninfl*, holding a critical role in granulocyte and macrophage-dependent immune responses and a critical role in NET formation (Liu *et al*, 2021). Upregulation of LGALS9, LRIG1, CXCL10, SIGLEC1, CD300C, and TCN2 in both *acute* phases compared to *Ctrl-noninfl* with significantly higher expression levels in the *acute-early* phase than in the *acute-late* phase pointed towards an innate immune response while involving factors that contribute to NET formation as well as stroke risk.

GLB1, OXT (log2(FC)>2), FGF21, IL34, BAG3, SEMA4C, TLR3, GPR37, ANXA5 were found to be significantly upregulated in the C19 intragroup comparison of disease phases, *i.e.,* during the *acute-early* disease course, whereas NCF2 was upregulated in the *acute-late* phase.

When comparing the *acute-early* to the *recovery* phase, we identified 59 differentially regulated proteins involved in innate and adaptive immunity, as well as remodeling processes with a predominant upregulation in the *acute-early phase*. The top 10 regulated proteins reflected this by the inclusion of inflammatory (CD300E, IL15, IFNG, LGALS9, CD14, CXCL10, IFNGR2, LILRB4), metabolic (CASC4), and remodeling processes (CDON9). Downregulated proteins were engaged in immune response mechanisms including cell adhesion, adaptive immune processes, cell-cell matrix interaction, and related metabolic activity (CD1C, TNFSF11, SELPLG, SKAP1 (inflammation), CDON, THBS4, MFAP5 (remodeling), PLTP (metabolism)) (**Figure 6A & B**, **Figure S3**).

Between the *acute-late* and the *recovery phase*, 17 proteins were found to be differentially regulated including inflammatory (BST2 (log2(FC)>2), SDC1, DDX58, LGALS9, LILRB4), and coagulation specific (C4BPB, ITIH3) processes together with *Remodeling of cardiac muscle and blood vessels* (CDH2, SMOC1) and other tissues (GFRA1, MDK, BAIAP2, SMOC1), and metabolic activity (CASC4, MME, FKBP5). Whereas these proteins were upregulated in the *acute-late* phase, the proteins ITGA11 and ROBO2 with a role in tissue remodeling were upregulated in the *recovery* phase (**Figure 6A & B**, bottom left circle). It has to be noted that important proteins such as BAIAP2 and GFRA1 that were identified by the phase comparison hold critical functions in the central nervous system.

While the majority of proteins, especially those associated with coagulation, showed a constant decrease in abundance over the disease trajectory, some proteins increased over time, *e.g.*, CD1C, SELPLG (inflammation), SKAP1, TNFSF11 (inflammation), PLTP (metabolism), CDON, and MFAP5 (remodeling). Other proteins displayed more complex regulation patterns such as delayed changes, e.g., ITGA11, ROBO2, and THBS4 remained unchanged in *acute* disease and increased in *recovery* phase, while ANGPTL1 and FGF21 decreased in the *acute-late* and FKBP5 in the *recovery* phase; or alternating patterns (e.g., PADI4, SDC1, and BAIAP2) (**Figure S4&S5**).

When comparing disease phase-specific regulation for both severity grades, disease phase-dependent regulation differed for *C19-ox* and *C19-nonox* patients in the *acute-late* phase of the disease. Here, the comparison showed an upregulation of 14 proteins in *C19-ox patients* indicating the activation of innate and adaptive immune response mechanisms, energy metabolism, stress response, and remodeling including modulation of growth factor signaling (CRNN, FOLR1, PAG1 (inflammation), CD177 (log2(FC)>2), HK2, MSLN, NOS1, SFTPA2, SRP14, TNFRSF10B (remodeling), PFKFB2, WFDC2, RASSF2, REG1A (metabolism)) (**Figure 6A & C, Figure S3**). No phase-specific regulated protein could be identified in *C19-ox* patients, while lower disease grades demonstrated differential regulation such as the upregulation of OXT (log2(FC)>2, metabolism) in the *acute-early* phase when compared to the *acute-late* phase together with the inflammatory markers IFNL1, IL6, BST2 (all log2(FC)>2), CD300E, LGALS9, and LILRB4 in comparison to the *recovery phase*. Additionally, coagulation and complement activation, e.g., upregulation of ITIH3 (coagulation) and C4BPB (coagulation) were observed in the *acute-late* phase when compared to the *recovery* phase. (**Figure 6A & D**, **Figure S3**).

Disease trajectory regulated inflammation associated pathways were primarily observed between *acute-early* and *recovery* phases, including interleukin signaling (*i.e*., *Interleukin-10 signaling*), *Neutrophil degranulation*, *Cargo concentration in the ER*, and *Immunoregulatory interactions between a Lymphoid and a non-Lymphoid cell*, capturing the innate and adaptive immune system, as well as *ROS regulation*. Remodeling processes further characterize the comparison between *acute-early* and *recovery* phase and comprised the *Regulation of Insulin-like Growth Factor transport and uptake by Insulin-like Growth Factor Binding Proteins*, as well as *Post-translational protein phosphorylation* via *Transport to the Golgi and subsequent modification*. *Transport to the Golgi and subsequent modification* was uniquely regulated in this comparison (**Figure 6F**).

For proteins contributing to NET formation, a gradual decrease was detected over time for CCL8, LGALS9, ANXA5, GRN, CTSC, and MME when comparing the *acute-early phase* to later stages, whereas PADI4 and NCF2 showed an increase in the *acute-late* phase (**Figure S4**), implicating neutrophil hyperactivation following the inflammatory peak.

**In summary,** the *acute-early* disease phase is specifically characterized by an innate immune, virus-related host response that involves vascular and matrix remodeling, while matrix remodeling proteins were also found to be upregulated in the *recovery* phase when compared to the *acute-late* phase (**Figure 6A & B & E & F, Figure S3)**. In contrast, the critical regulator of NET formation PADI4 was differentially regulated in the *acute-late* phase. Protein expression pattern in both acute phases indicated regulation of innate immune defense mechanisms such as activation and recruitment of leukocytes, autophagy and indicated by the regulation of CXCL10, SIGLEC1, CD300C, NCF2, ANAX5, and BAG3 and matrix remodeling as identified through the differential expression of SEMA4, LGALS9, and GLB1, promoting mesenchymal activation and matrix formation. Interestingly, TCN2, engaged in vitamin B12 uptake, has been described to modify stroke risk (Hsu *et al*, 2011), whereas GPR37 signaling has been shown to modulate the migration of olfactory ensheathing cells (Saadi *et al*, 2019). This expression pattern was most prominent in *C19-ox* patients who showed a strong time-dependent regulation of innate, adaptive immune, and stress response, as well as activation of the coagulation and complement system, *e.g.*, upregulation of ITIH3 (coagulation) and C4BPB (coagulation), in the *acute-late* phase when compared against the *recovery* phase (**Figure 6A** **& C & D, Figure S3**).

## Discussion

Although to date, numerous studies have described the wide range of symptoms of severe SARS-CoV-2 infection, e.g., acute respiratory distress syndrome (ARDS), lymphopenia, coagulopathy, and multi-organ damage (Bernardes *et al*, 2020; Faust *et al*, 2020; Wiersinga *et al*, 2020), a detailed analysis of the underlying sequence of events is still missing. Studies that targeted protein regulation in COVID-19 patients aimed for a better understanding of disease-related processes while trying to identify potential biomarkers at the same time and have reported different immune response-related phenomena. The so-called “cytokine storm” comprised regulation of CXCL8, CXCL10, IL-6, TNFalpha, and IFNG, indicating that the synergism between TNF-α and IFNG, known to trigger inflammatory cell death and tissue damage, may account for SARS-CoV-2 mortality due to cytokine shock (Karki *et al*, 2021; Buszko *et al*, 2021; Yang *et al*, 2021) and potentially addressed by existing therapies (Tang *et al*, 2020).

Our study addressed the gap of existing knowledge with regard to a differentiated understanding of disease dynamics while specifically considering disease stage and severity, thereby significantly adding to existing knowledge in the field (**Figure 7**). Rooting the protein markers detected by an unbiased approach in disease pathophysiology, we achieved the identification of critical disease-stage and -phase-specific indicators in high-risk COVID-19 patients. We both confirmed as well as newly discovered urgently needed markers in a COVID-19 patient population that is omnipresent in university hospitals due to diverse preexisting conditions.

**Figure 7:**
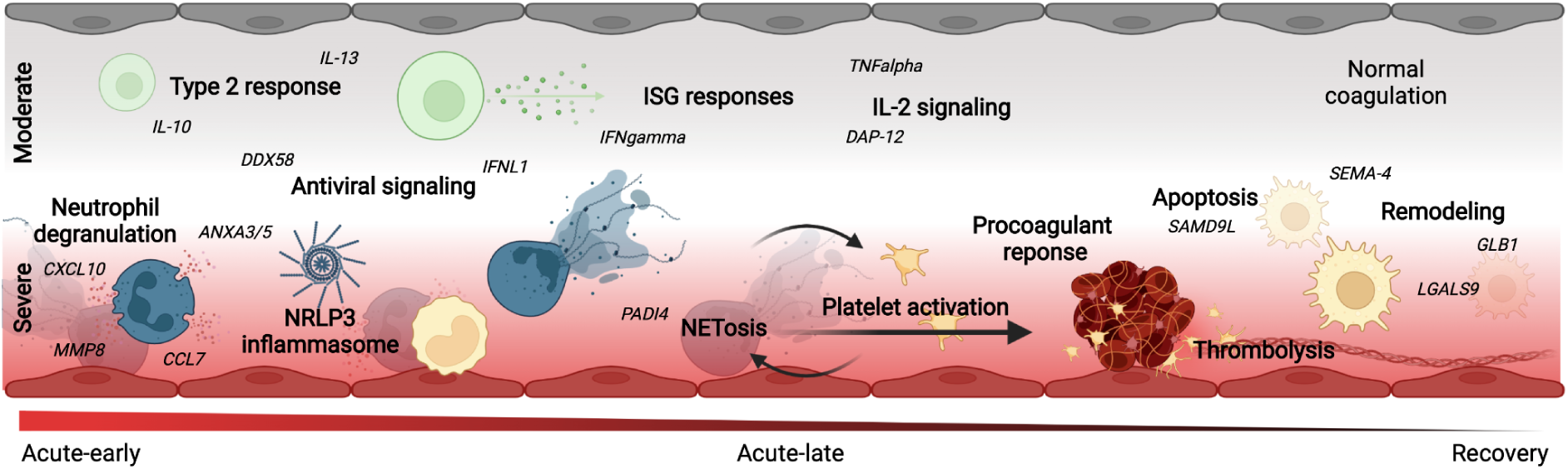
Schematic overview of the disease-phase and disease-severity-dependent pathophysiologic response in moderate (WHO≤4, upper half, *C19-nonox*) and severe (WHO≥4, lower half, *C19-ox*) *C19* patients. While sharing a significant pattern of protein regulation, *i.e.*, TNF signaling, severe disease was characterized by antiviral- and NETosis-related protein regulation including coagulation activation. Moderate disease associated patterns were dominated by a type-2 immune response and IL-2 associated signaling. Both disease severity grades progress from acute inflammation to the activation of remodeling processes.

To relate the plasma protein signatures detected by proteome screening to clinically relevant disease phases while considering internationally accepted disease severity grades, we studied COVID-19 patients in comparison to both non-inflammatory and inflammatory control subjects. To sensitively address the heterogeneous disease characteristics in a multimorbid patient cohort, we improved disease phase assignment by defining novel individual clinical trajectories using the inflammation markers IL-6 [pg/ml] and CRP [mg/dl]. In contrast to our approach, previous SARS-CoV-2 studies solely relied on disease phase assignment relating to the number of days after symptom onset, PCR result, or hospital admission, while other studies primarily referred to disease symptomatology (Schulte-Schrepping *et al*, 2020; Lam *et al*, 2021). In general, cutoffs of inflammation markers have been used with good success in predicting COVID-19 severity at admission (Herold *et al*, 2020) but did not consider individual trajectories and threshold of laboratory variables that are likely of importance when studying a cohort with significant preexisting conditions and related medications.

The significant variation in the course of the disease when comparing different patient groups and treatment settings (Tolossa *et al*, 2021), including the average time for symptom resolution (2 to 71 days (Abrahim *et al*, 2020) or 10-14 (mild disease) to 21-42 (severe disease) days (Bhapkar *et al*, 2020; Jin *et al*, 2020)), likely renders solely ‘time-after-infection’ based disease phase assignment inaccurate, especially in patients with multiple influencing factors. In contrast to previous approaches (Schulte-Schrepping *et al*, 2020), we, however, demonstrated that the trajectories based on the individual course of critical routine laboratory markers, more adequately matched clinical symptoms and critical events in the patient cohort studied, resulting in better discriminatory power for (severity-dependent) disease phase separation.

We showed that the inflammatory response in *C19* patients obtained by the comparison to healthy individuals is characterized by a strong induction of innate immune response mechanisms in the *early-acute* phase as indicated by the regulation of *TLR through endogenous ligands*, *Neutrophil degranulation together with cargo concentration in the ER,* and *COPII-mediated vesicle transport* accompanied by the parallel upregulation of prothrombotic and thrombolytic phenomena, *i.e.*, *Dissolution of Fibrin Clot*, *Response to elevated platelet cytosolic calcium*, and *Platelet degranulation*. The significant overlap of this protein signature with the disease-specific response revealed by the comparison to patients with signs of non-SARS-CoV-2 related inflammation the importance of interferon related antiviral signaling, i.e., DDX58 as confirmed by previous studies (Winheim *et al*, 2021), together with the regulation of apoptosis and DNA damage, accompanied by the activation of TLR signaling, lysosomal functions and an indication of epithelial cell damage.

This response could be largely attributed to severe disease, whereas protein regulation in mild-to-moderately affected *C19* patients was dominated by the disease-stage specific regulation of interleukin-2 and INFG signaling, as well as the shared regulation of remodeling processes as indicated by the regulation of *Extracellular matrix organization*, *Post-translational protein phosphorylation,* and *Regulation of Insulin-like Growth Factor transport and uptake by Insulin-like Growth Factor Binding Proteins* in the *acute-early* and *late* phase. However, the differential regulation indicated no activation of coagulation processes in less diseased patients in comparison to the *Ctr-noninfl* group although anticoagulation treatments were equally administered in both groups (*C19-nonox*: 62.16%, *C19-ox*: 59.26%). Interpretation of these findings, however, needs to take into account that deaths in the *C19-ox* group resulted in the overrepresentation of survival-related changes to protein expression in later disease phases.

The strong upregulation of proteins related to NET formation (Wang *et al*, 2020; Brinkmann *et al*, 2004) was observed during both *acute* COVID phases (*acute-early*: 58/78, *acute-late*: 54/78) with 36 proteins similarly regulated. Activation of NET formation in context with other indicators of inflammasome activation (Bardoel *et al*, 2014; Middleton *et al*, 2020; Nicolai *et al*, 2020; Zheng *et al*, 2020; Zhou *et al*, 2021; Smet *et al*, 2021) specifically characterized patients with severe disease (WHO >= 4). The regulated proteins included CD177, a prominent activation marker present on the surface of circulating neutrophils (Nicolai *et al*, 2020; Bai *et al*, 2017), MME (CD10) as an immaturity marker of neutrophils and previously associated with severe COVID-19 (Schulte-Schrepping *et al*, 2020; Kaiser *et al*, 2021), PDGFRA as a marker of platelet degranulation, and PADI4 as a key regulator of NETosis, whereas the classical NETosis/degranulation marker MPO was not found to be regulated in any of the comparisons. Similarly, strong activation of inflammasome related processes was indicated by the regulation of AGER as an important regulator of CASP-11 inflammasome activation (Chen *et al*, 2019a) side-by-side with an upregulation of IL-1 and IL-18 (Zheng *et al*, 2020), as well as IL-6 and TNF expression together with an overenrichement of *TNF receptor superfamily (TNFSF) members mediating non-canonical NF-kB pathway (Zheng et al, 2020)*. Regulation of PLAUR points towards thromboembolic phenomena in these patients (Nicolai *et al*, 2020; Schulte-Schrepping *et al*, 2020; Wilk *et al*, 2020), which were controversially discussed for their dependence on disease severity (Nicolai *et al*, 2020).

When tracking the disease course, we observed the differential regulation of protein expression related to angiotensinogen, surfactant and SIRP metabolism, ROS regulation, and IL-6 signaling during early disease in the overall comparison and especially in the *C19-ox* patients in comparison to the non-inflammatory control group, whereas the later phase is characterized by the predominant regulation of proteins associated with matrix degradation and apoptosis.

On the one hand, we hereby show regulation of significant players in the immune host response confirming the role of inflammatory cell death and tissue damage (Karki *et al*, 2021). On the other hand, we were able to add to previous studies by showing the dynamic of NETosis and inflammasome regulation (Bardoel *et al*, 2014; Middleton *et al*, 2020; Nicolai *et al*, 2020; Zhou *et al*, 2021; Smet *et al*, 2021) in severely affected patients in contrast to a type-2 centered immune response involving interleukin 4, 10, 13, and TNF signaling in both disease groups or in less severe disease only (*e.g.*, *Interleukin-2 family* and *IN signaling*). These changes were found to be accompanied by remodeling processes.

Activation of the coagulation system was primarily detected in severely diseased patients in our cohort, although clinical reports also detected thromboembolic events in less severe disease (Chen *et al*, 2021; Clavijo *et al*, 2021), potentially due to the lack of detection regarding local, organ-specific events. Activation of the coagulation system in more severely diseased patients, as well as activation of the complement system likely drives thrombo-inflammation in COVID-19 (Afzali et al. 2021).

Regulation of proteins such as CA6 (associated with hypogeusia) or TCN2 (associated with stroke) identify disease characteristics, thereby supporting the significant potential of our unbiased approach to inform both pathophysiologic understanding and biomarker development.

Previous studies that employed proteome analysis mirrored our findings such as activation of the complement system, monocyte signaling (CD14, proteins of the LGAL family) and inflammation (CD48, SIRPB1) (Park *et al*, 2020), as well as the regulation of different plasma protease inhibitors such as ITIH3 (Geyer *et al*, 2021; Messner *et al*, 2020; Park *et al*, 2020; Shen *et al*, 2020) in COVID-19. Further in line with our findings, vascular markers such as vWF and proteins indicating coagulation activation were found to be regulated in previous studies, but in contrast to our studies described an early decrease (Messner *et al*, 2020; Shen *et al*, 2020). Likewise, proteins involved in metabolic processes such as lipoprotein homeostasis (PLTP) were differentially regulated in COVID-19 patients.

Enabling us to put the proteomic signatures into perspective and validate disease phase assignment, we comprehensively tracked biochemical indices. Here, comparable changes were observed in *C19-ox* and non-C19 related inflammation (*Ctrl-infl*) patients including, despite its common use in SARS-CoV2 (Liu *et al*, 2020a), nondiscriminatory CRP levels when comparing C19-ox patients with subjects suffering from non-COVID related inflammation. Proteomic analysis, however, significantly broadened the picture by demonstrating significant differences in protein expression between these groups. In contrast, PTT levels differed between *C19-ox* and *Ctrl-infl* patients, in line with the observed proteomic pattern indicating coagulation activation in *C19-ox* patients. Although shortened PTT times were discussed to predict poor outcome in patients with varying diseases (Reddy *et al*, 1999), its role in COVID-19 is still controversially discussed (Devreese, 2021). Changes in proteome pattern during the course of disease in each severity group were mirrored by cell numbers as well as coagulation and inflammation markers.

Regarding differential blood counts, the analysis in *C19-ox* patients indicated lymphopenia, low monocyte levels, and neutrophilia when compared to *C19-nonox* patients, in line with previous studies (Huang *et al*, 2020; Williamson *et al*, 2020; Lombardi *et al*, 2020) but again did not differentiate well severely affected patients from subjects with inflammation of different origin (*Ctrl-infl*) supporting the controversial discussion of their predictive value (Woodruff *et al*, 2020). In context with the changes in differentiated blood cell counts, the proteome changes likely reflected the activation of the immune system and again indicated the added value in delineating COVID-19 immune responses in relation to disease severity and -phase (**Figure 3 & 4**). Lower monocyte levels in more severely affected patients, however, could relate to the extravasation of these cells and suggest the subsequent activation of macrophages as indicated by the observed proteomic signature (Arango Duque & Descoteaux, 2014).

Association of INFG and TNF-associated monocyte polarization and the pro-fibrotic potential of monocyte-derived alveolar macrophages underline the potential of the observed signature to induce long-term remodeling (Castro *et al*, 2018; Misharin *et al*, 2017).

With regard to the impact of (co)morbidities, previous studies identified risk factors that were in part reflected in our cohort. Whereas *C19-ox patients* were characterized by increased age when compared to *C19-nonox* patients in line with previous studies (Huang *et al*, 2020; Williamson *et al*, 2020), we could not observe a higher rate of male patients in more severe disease (Huang *et al*, 2020; Williamson *et al*, 2020). Similarly, we did not observe a significantly reduced time between symptom onset and hospitalization for more diseased patients (Liu *et al*, 2020b; Zhou *et al*, 2020), but confirmed a longer hospital stay (Liu *et al*, 2020b; Zhou *et al*, 2020), higher rates for ICU admission (Liu *et al*, 2020b; Zhou *et al*, 2020; Grasselli *et al*, 2020), invasive ventilation (Liu *et al*, 2020b; Zhou *et al*, 2020), and adverse outcome. Further, we demonstrated a higher incidence of comorbidities, e.g., pre-existing lung disease, e.g., COPD or asthma (Huang *et al*, 2020; Williamson *et al*, 2020), hypertension (Huang *et al*, 2020; Williamson *et al*, 2020), diabetes (Huang *et al*, 2020; Williamson *et al*, 2020), kidney diseases (Williamson *et al*, 2020), or impaired immune function (Huang *et al*, 2020; Williamson *et al*, 2020) in more diseases patients (*C19-ox*). In contrast, we did not find disease symptoms more prevalent in *C19-ox* patients compared to *C19-nonox* patients with the exception of fatigue, dyspnea, and an increased incidence in secondary infections, in line with previous studies (Liu *et al*, 2020b; Zhou *et al*, 2020) and in part explaining the increased duration in hospital stay.

Limitations of the present study include its observational design and the retrospective analysis resulting in missing data in a small number of patients. Partially counteracting these imitations, the study benefits from homogenous and comprehensive clinical monitoring in a high-risk patient collective that continuously dominates patient admission in university hospitals during the COVID pandemic. While providing a very good basis for biomarker identification in different disease phases and severity grades, results have to be confirmed in targeted approaches in different clinical centers. These prospective studies need to include - amongst others - environmental or social factors not investigated in the current study while considering the impact of emerging SARS-CoV-2 variants and the effect of the potentially gender-dependent vaccination status, not present in the first pandemic wave addressed in our approach (Ovies *et al*, 2021).

*In summary,* we identified a COVID-related protein signature that indicates an antiviral response together with NET / inflammasome activation predominantly driven by their regulation in severely affected patients. In contrast, regulation in less severely diseased patients was found to be characterized by a type-2 centered immune response. The findings were enabled by the newly identified disease trajectories based on the individual course of important routine laboratory variables. This approach both confirms findings from previous studies and also facilitates the identification of new proteins with significant potential to serve as COVID-19 disease indicators at the same time.

## Materials and Methods

### Clinical Data Collection, patient grouping, and disease phase assignment

The study prospectively enrolled 65 patients with PCR confirmed SARS-CoV-2 infection during the first phase of the COVID-19 pandemic in Germany (03/2020 to 08/2020), before steroid treatment for SARS-CoV-2 was routinely prescribed. Patients were enrolled shortly before or at the onset of the acute infection phase when laboratory signs of infection and disease-specific symptoms develop. Twenty-five patients with acute (inflammatory control group; *Ctrl-infl*) or no/low non-COVID-19 related inflammation (healthy control group; *Ctrl-noninfl*) were additionally included in the study as control groups. Patients and control subjects are part of the COVID-19 Registry of the LMU University Hospital Munich (CORKUM, WHO trial id DRKS00021225). Patient data were anonymized for analysis. The study was approved by the ethics committee of the Ludwig-Maximilians-Universität (LMU), Munich, Germany (Study title: “COVID-19 Register des LMU Klinikums (CORKUM)”; Project No: 20-245 (initial approval date: 03/2020; Amendment approval dates: 07/2020, 01/2021, 05/2021) as well as under the Project No: 20-259 (CPC-M bioArchive)).

Comprehensive electronic health records of all 90 patients were provided including baseline information like age, gender, medical background about comorbidities, and medication before hospital admission. Furthermore, information about the clinical course was provided including different routine biomedical indices (*e.g.*, blood cell measurements), different inflammation markers like CRP, IL-6, or Ferritin, coagulation markers such as platelet count or PTT, and other body function values (*e.g.*, creatinine or hsTroponinT, measured at admission as well as repeatedly over the hospital stay as needed), received treatments (*e.g.*, ventilation or medication), and adverse events (*e.g.*, acute kidney failure, thrombosis/embolism, or death).

COVID-19 patients were classified according to ordinal scale for clinical improvement of COVID-19 infection reported by the WHO (Blueprint, 2020) and grouped into two sub-cohorts based on the need for oxygen supply, *i.e*., disease severity (WHO≥4 - *C19-ox*; WHO≤3 - *C19-nonox*) (**Figure 1A**).

To specify the host immune response to COVID-19 infection while considering the underlying disease phase, we developed a novel approach for a high-risk, multimorbid patient cohort to improve upon general time-of-infection based approaches (e.g., (Schulte-Schrepping *et al*, 2020)) or approaches using thresholds for levels of inflammation markers (*e.g.*, (Herold *et al*, 2020; Chen *et al*, 2020a)). As IL-6 (> 80 pg/mL) and CRP levels (> 97 mg/L) correctly classified 80% of patients regarding their risk of respiratory failure (Herold *et al*, 2020), we used these markers and extended this approach by defining disease phases based on individual trajectories while considering important clinical hallmarks. Using inflammation markers, we individually identified an inflammation peak for each patient, defined as the time point of the highest measured CRP or IL-6 value (whichever occurred later) broadened by a window of 24h after this peak to account for individual differences in inflammation marker decline. Accordingly, a total of three disease phases were distinguished: *COVID-19-acute-early*, *COVID-19-acute-late*, and *COVID-19-recovery phase*. The *COVID-19-acute-early phase* was defined as the time between disease onset, *i.e.*, the onset of clinical symptoms and/or first positive PCR test and the end of the inflammation peak. The *COVID-19-acute-late phase* was defined as the time after the inflammation peak until hospital discharge. The *COVID-19-recovery phase* was defined as the time after hospital discharge.

In cases with significant discrepancy of disease severity, *i.e.*, maximum WHO score at admission and the individual trajectory of IL-6 and CRP, the samples were assigned to the *acute-late phase*, assuming a surpassed inflammation peak at admission **(Figure 1B, Figure S1**). Three samples (Patient 12 and 18, **Figure S1**) without any IL-6 and CRP peaks were assigned based on the proteomic data, by applying a k-nearest neighbors clustering algorithm to assign the samples to their most likely disease phase while validating clinical symptoms for group assignment.

*Non-C19* patients were assigned to two control groups based on the presence of inflammation: We included 14 patients with acute non-COVID related inflammation (*Ctrl-infl*) characterized by a maximum CRP>0.5 mg/dl or IL-6>5.9 pg/ml and 11 subjects without elevated inflammation markers (*Ctrl-noninfl*), *i.e.*, CRP≤0.5 mg/dl and IL-6≤5.9 pg/ml (**Figure 1A**). Phase and group definitions can be found in **Table 1**.

### Sample Collection and Processing

A total of 129 samples were collected in all study subjects with one to five serial samples over the different phases and subjected to proteomic analysis. Plasma was separated from heparinized whole blood by centrifugation at 2,000 g for 15 minutes at room temperature and immediately stored at -80° C until preparation for proteome analysis.

### Olink plasma proteomics

The Olink® Explore 1536 platform was used to measure protein abundance in plasma samples. The full library consisting of four 384-plex panels (Inflammation, Oncology, Cardiometabolic, and Neurology) was used to screen 1,472 proteins. Relative protein abundance was calculated from the number of matched counts on the Illumina NovaSeq 6000 run using two S1 flow cells with 2 × 50 base read lengths. The counts of protein specific-barcode sequences were transformed into Normalized Protein eXpression (NPX) units and an intensity normalization algorithm was applied to reduce the technical variation. The final data were provided in the arbitrary unit (NPX) on a log2 scale. Quality control (QC) was performed at both protein and sample levels. Three internal controls are spiked into each sample in order to monitor the quality of assay performance, as well as the quality of individual samples. Following criteria are applied to pass the sample QC: the average matched counts for each sample must exceed 500 counts; the deviation from the median value of the incubation- and amplification controls for each sample should not exceed +/-0.3 NPX for either of the internal controls. We, therefore, excluded 8 samples, whose mean of the failing proteins deviated more than 0.5 standard deviations from the overall mean of all samples which passed QC. As a further QC instance for comparability, the three proteins TNF, CXCL8, and IL-6 were measured in each of the four Olink® Explore panels. Since all four measurements were highly correlated for each of the three proteins (TNF: r=0.952-0.965; CXCL8: r=0.989-0.998; IL-6: r=0.979-0.997), we kept only one representative for each protein based on the minimal number of QC warnings, conformity in scatter plots, and population variances. Furthermore, Olink® recommends that proteins with a large proportion of samples below the limit of detection (LOD) should be excluded from the analysis. We, therefore, excluded 77 proteins that were under the LOD in more than 25% of samples in all study groups.

### Data analysis

We compared clinical covariates and routine biochemical indices within the first 24 hours after hospital admission separately for *C19-ox* and *C19-nonox* with both control groups (*Ctrl-infl* and *Ctrl-noninfl*). In addition, we analyzed routine biochemical indices and plasma proteomics by (1) comparing each phase (*acute-early*, *acute-late*, *recovery*) separately with both control groups *(Ctrl-infl* and *Ctrl-noninfl*) - overall (*i.e.*, severity-independent), and within *C19-ox* and *C19-nonox*; (2) comparing the phases (*acute-early*, *acute-late*, *recovery*) with each other - overall (severity-independent) and within *C19-ox* and *C19-nonox*; and (3) comparing the two severity groups (*C19-ox* and *C19-nonox*) with each other in each phase (*acute-early*, *acute-late*, *recovery*).

Continuous and categorical variables were presented as median (interquartile range (IQR) with 25% and 75% percentiles) and *n* (%) respectively. We used the Mann-Whitney-Wilcoxon test, Welch test, Tukey’s range test, X^2^ test, and Dirichlet regression to compare differences between the C19 groups and the *Ctrl-infl* / *Ctrl-noninfl* control groups where appropriate. All tests were two-sided, and a P-value less than 0.05 was considered statistically significant. We used Python’s SciPy package (Inglett *et al*, 2015) to perform the statistical analysis. The effect sizes are described as log2 fold change (FC).

For the proteomics analysis, we used the R package limma (“Linear models for microarray data”) (Ritchie *et al*, 2015) adjusted for the following confounders: age, gender, cardiovascular diseases, diabetes, high cholesterol, lung disease, kidney disease, immuno-compromised status, superinfection during proteomics sampling, steroid treatment during hospital stay during or before proteomics sampling (**Table S11**). Volcano Plots were created using the R package EnhancedVolcano (Blighe *et al*, 2019). We conducted overrepresentation tests (based on hypergeometric models with a minimum count of three proteins) for biological processes and pathways using ClusterProfiler (Yu *et al*, 2012) and ReactomePA (Yu & He, 2016), while the Enrichplot (Yu, 2018) package was used for visualization of the overrepresentation results. All tests for the proteomics analysis were corrected for multiple testing using Benjamini-Hochberg correction, where a P-value less than 0.05 was considered statistically significant.

For the significance tests, one sample per person was used in order to avoid autocorrelation. For the *acute-early* and *acute-late phases,* we used the proteomics sample, which was collected closest in time to the median difference to the inflammation peak in that phase. For comparisons with the *recovery phase*, we always used the very last sample, no matter which group it was compared to, in order to keep the sample size as large as possible. In the non-C19 group, only one person had serial samples. We used the sample which was closest to the median CRP value of the non-C19 group.

We chose the clinical sampling closest to the proteomics sampling and accepted a range of four days (**Figure S6**). Not all biochemical indices were available at any given time point (**Figure S7**). Furthermore, no D-Dimer and IL-6 values were available in the *Ctrl-noninfl* group.

## Supporting information

Supplement Overview

## Data Availability

The data that support the findings of this study are available from the corresponding authors upon request. Access to data for research purposes is possible for eligible institutes according to GDPR rules.

## Acknowledgments

We would like to thank all CORKUM investigators and staff. The authors thank the patients and their families for their participation in the CORKUM registry.

## Author contribution

**Data Curation:** Johannes C. Helmuth, Clemens Scherer, Maximilian Muenchhoff, **Project administration**: Benjamin Schubert and Anne Hilgendorff; **Funding acquisition**: Stefanie M. Hauck, Jürgen Behr, Li Deng, Narges Ahmidi, Benjamin Schubert, Anne Hilgendorff; **Investigation**: Johannes C. Helmuth, Clemens Scherer, Maximilian Muenchhoff, Rainer Kaiser; **Methodology:** Alina Bauer, Elisabeth Pachl, Benjamin Schubert.; **Resources**: Johannes C. Hellmuth, Clemens Scherer, Maximilian Muenchhoff, Marion Frankenberger, Stefanie M. Hauck, Daniel Teupser, Nikolaus Kneidinger, Hans Stubbe, Agnese Petrera; **Software**: Alina Bauer and Elisabeth Pachl; **Supervision**: Narges Ahmidi, Benjamin Schubert, Anne Hilgendorff; **Validation**: Benjamin Schubert, Anne Higendorff, Jürgen Behr, Rainer Kaiser, Daniel Teupser; Bernhard Ryffe; **Visualization**: Alina Bauer and Elisabeth Pachl; **Writing - original draft**: Alina Bauer and Elisabeth Pachl; **Writing - review and editing**: Benjamin Schubert and Anne Hilgendorff; All authors commented on previous versions of the manuscript. All authors read and approved the final manuscript.

