## Supplement Overview for "Proteome reveals antiviral host response and NETosis during acute COVID-19 in high-risk patients"

#### Supplementary Tables

Table S1: Clinical characteristics for different subgroups of the cohort at admission

Table S2: Results of subgroup comparisons for clinical characteristics and biomedical indices at admission

Table S3: Details of statistical analysis of clinical parameters

Table S4: Reference values for routine biochemical indices

Table S5: Routine biochemical indices for disease phase assignment

Table S6: Log2(fold change) of routine biochemical indices for groupwise comparisons

Table S7: Differential Abundance Analysis Results: Phases compared to control groups

Table S8: Pathway Enrichment Analysis: Phases compared to control groups

Table S9: Differential Abundance Analysis Results: Trajectory comparisons

Table S10: Pathway Enrichment Analysis Results: Trajectory comparisons

Table S11: Description of confounders

### Supplemental Figures

Figure S1: Individual patient trajectories of CRP and IL-6

Figure S2: Severity-based trajectory of routine biochemical indices

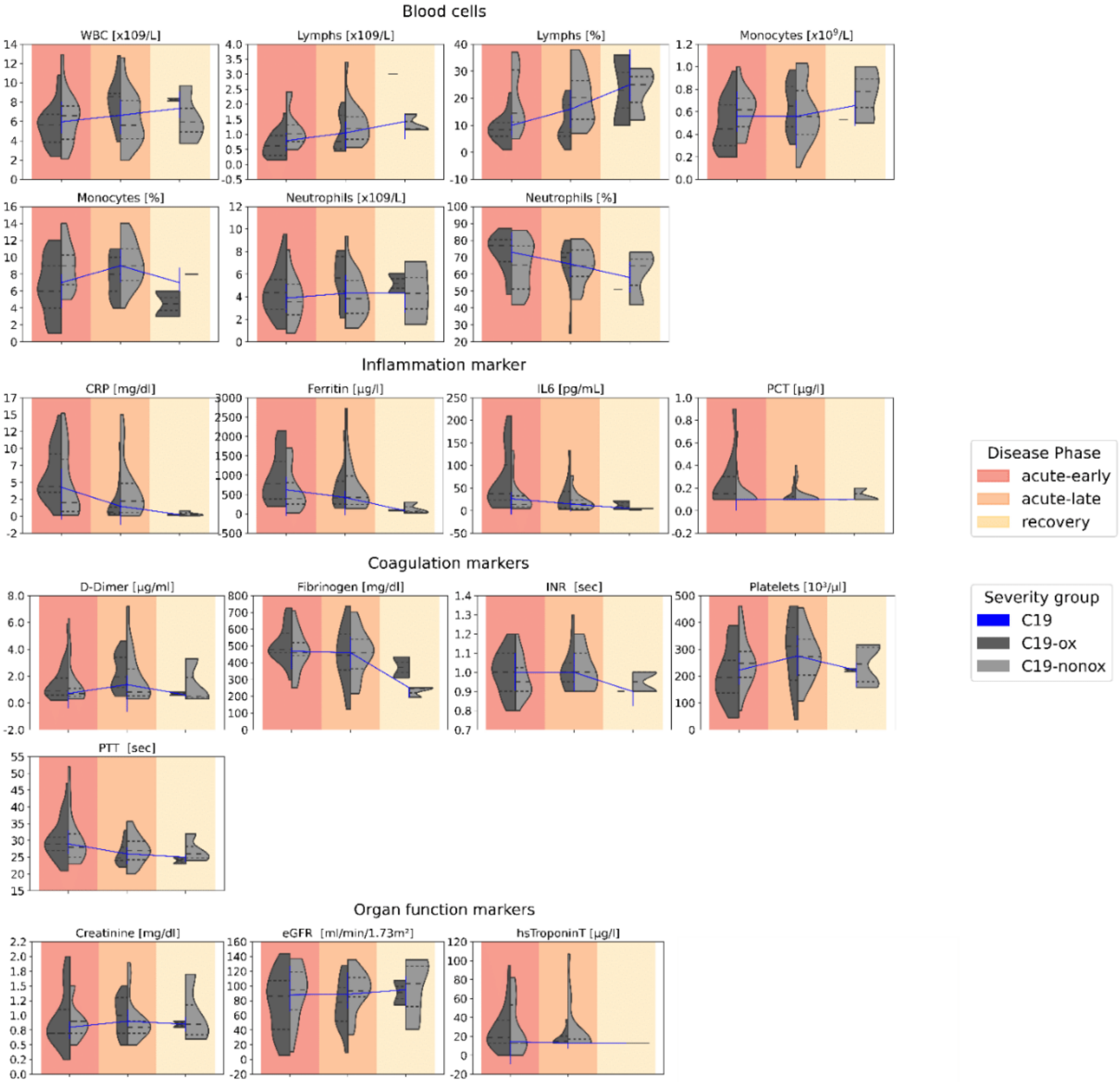

### Figure S3: Enrichment Analysis for Comparisons from Figure 4

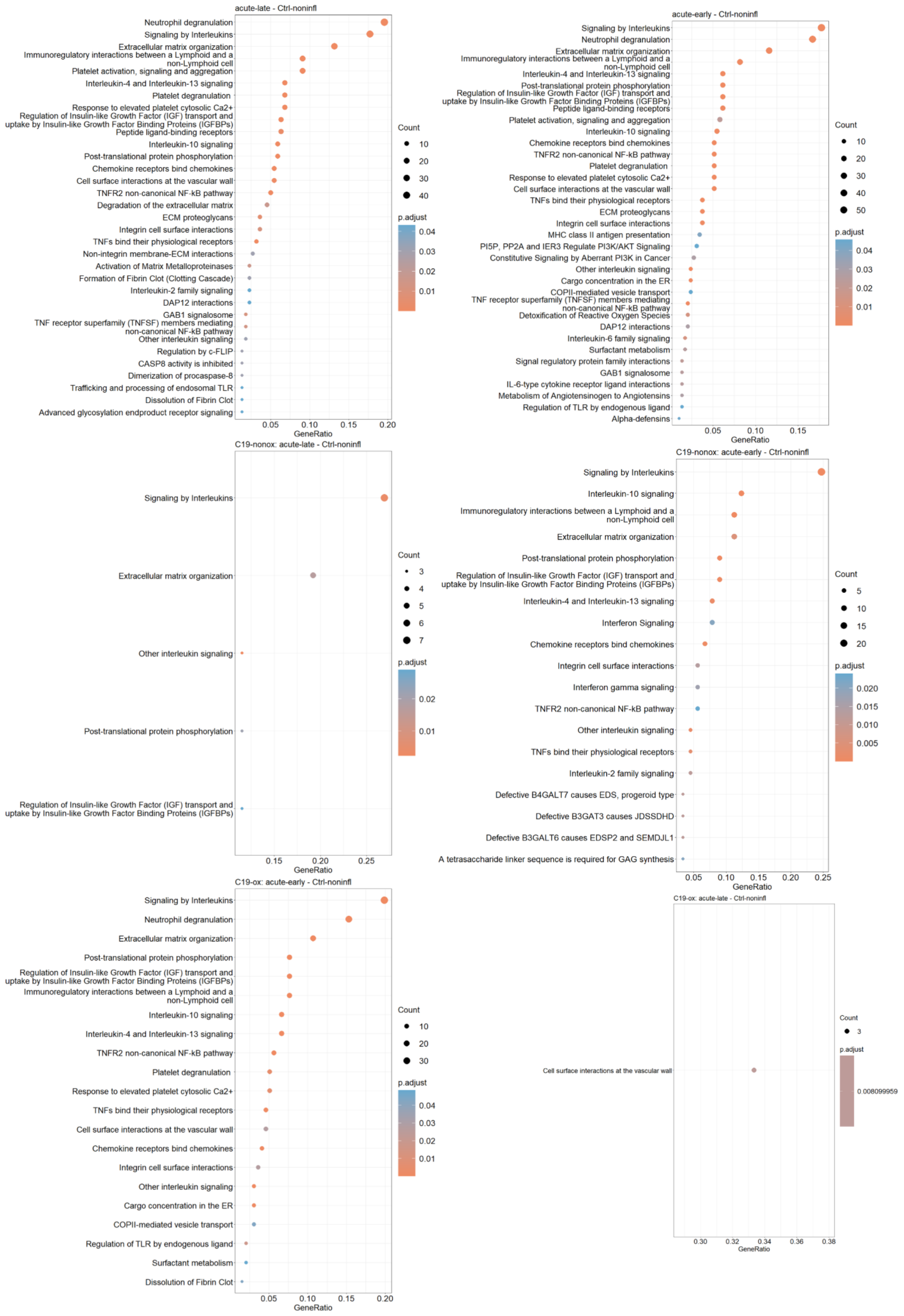

### Figure S4: Volcano plots and Enrichment Analysis from Figure 6

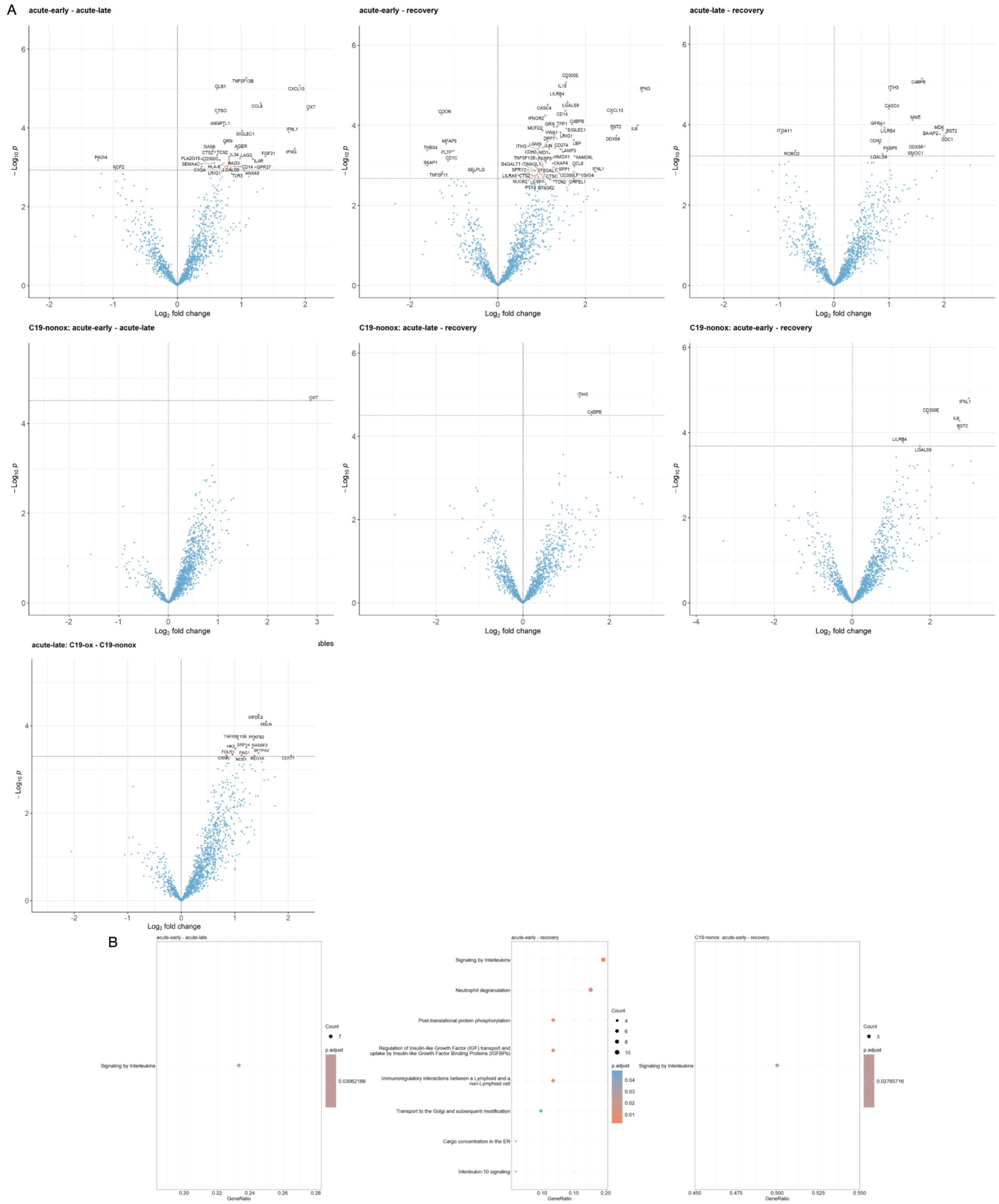

Figure S5: Boxplot for Comparisons

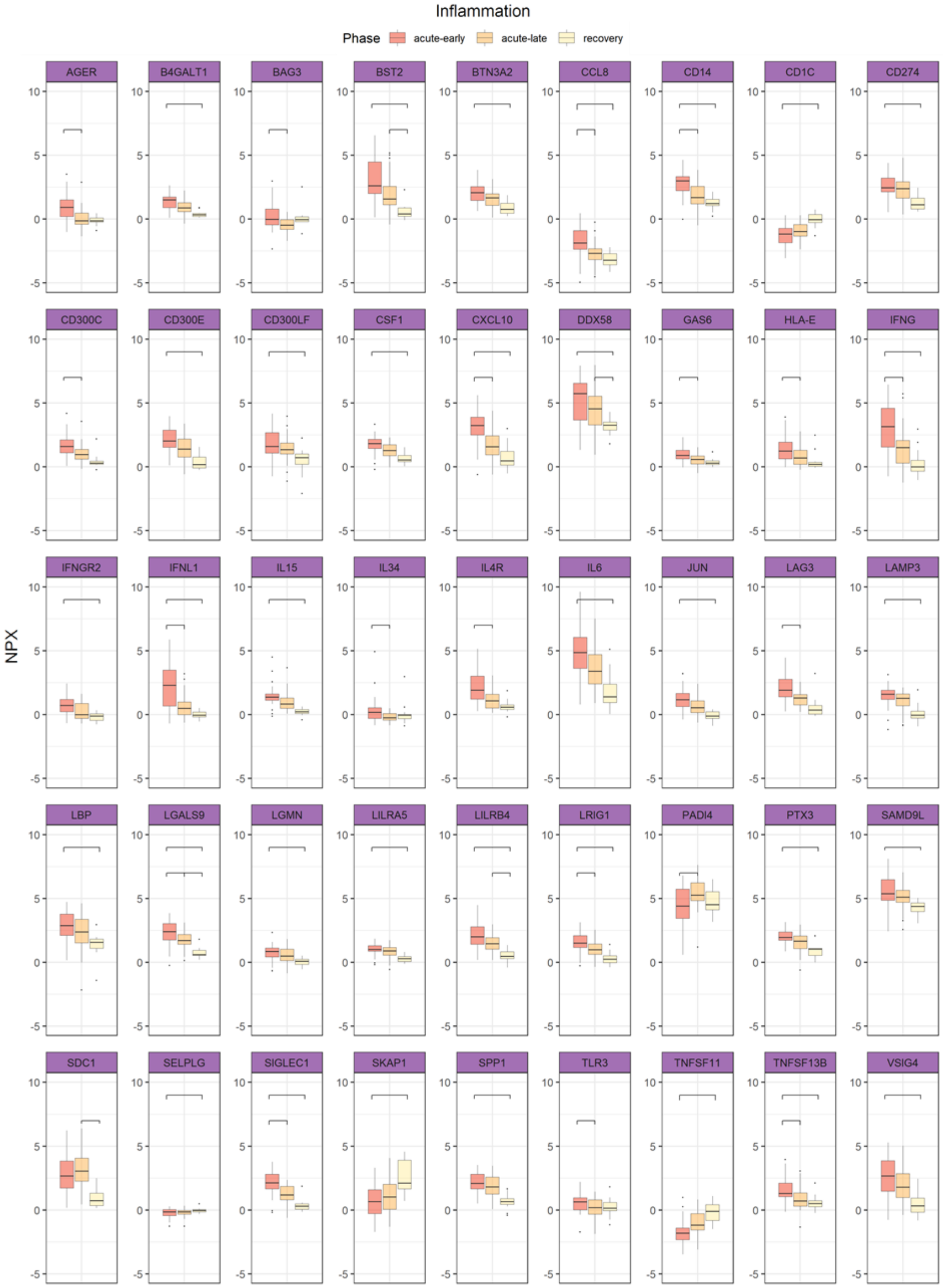


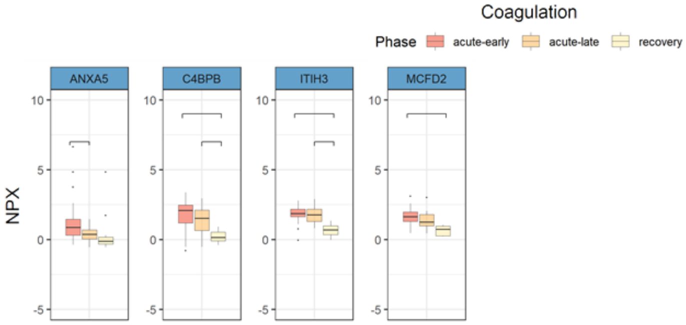

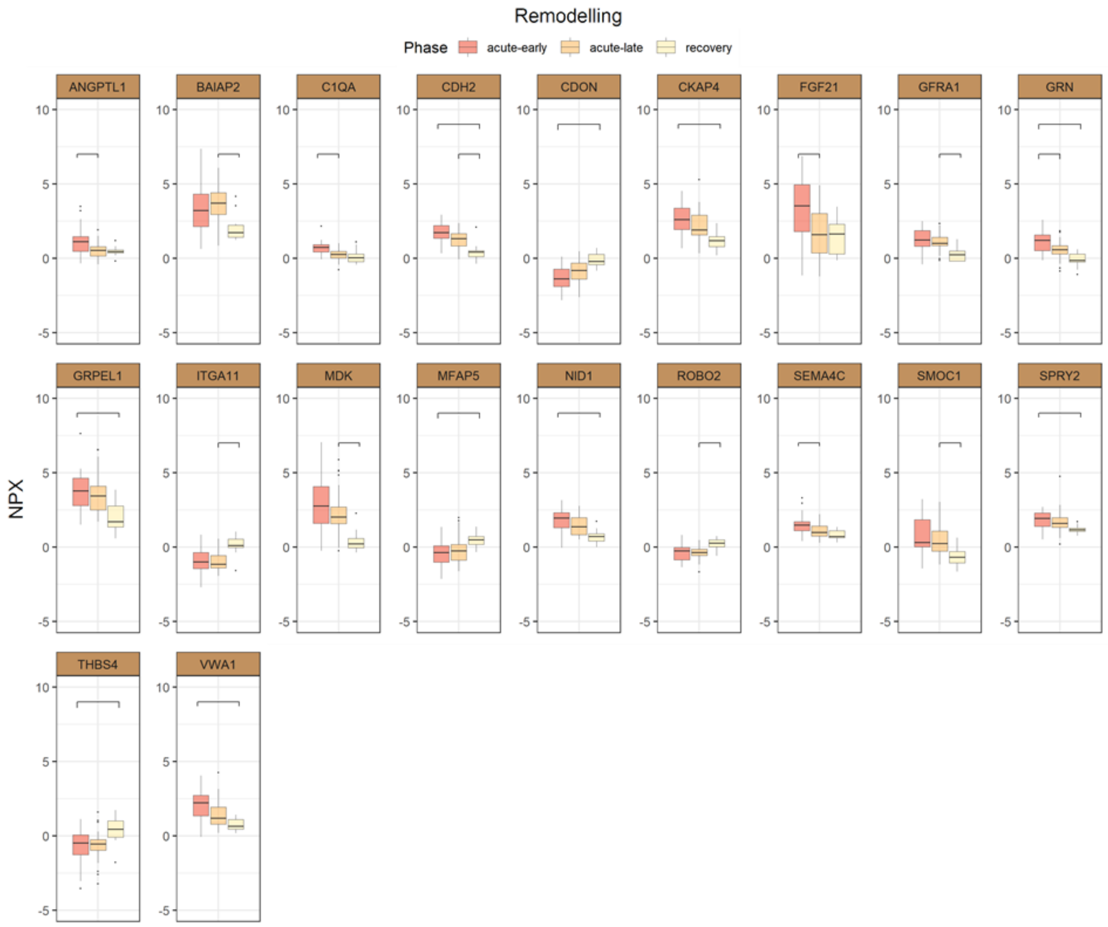
Remodelling
Phase
acute-early
acute-late
recovery
ANGPTL1
BAIAP2
C1QA
CDH2
CDON
CKAP4
FGF21
GFRA1
GRN
GRPEL1
ITGA11
MDK
MFAP5
NID1
ROBO2
SEMA4C
SMOC1
SPRY2
THBS4
VWA1
NPX
10
5
0
-5

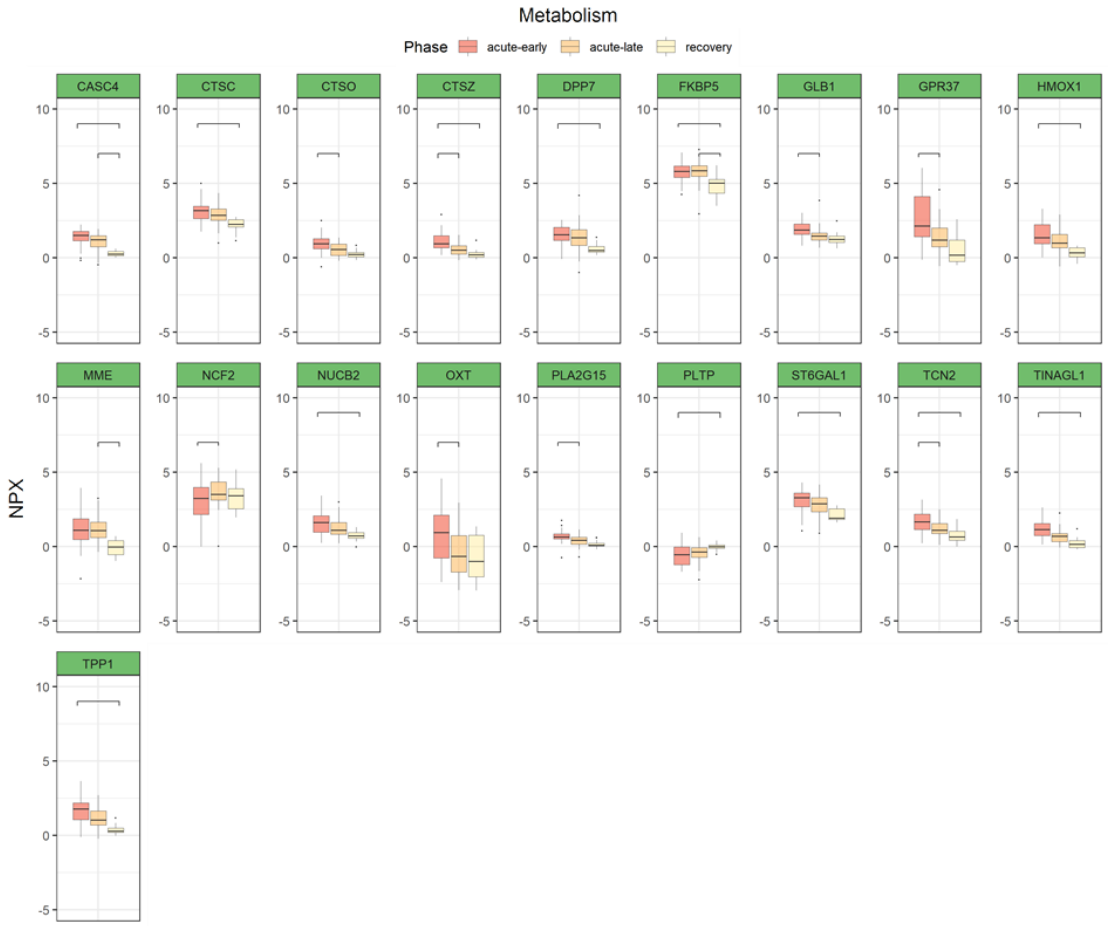
Metabolism
Phase
acute-early
acute-late
recovery
CASC4
CTSC
CTSO
CTSZ
DPP7
FKBP5
GLB1
GPR37
HMOX1
MME
NCF2
NUCB2
OXT
PLA2G15
PLTP
ST6GAL1
TCN2
TINAGL1
TPP1
NPX
10
5
0
-5

#### Figure S6: Time between nearest laboratory sample and protein sample

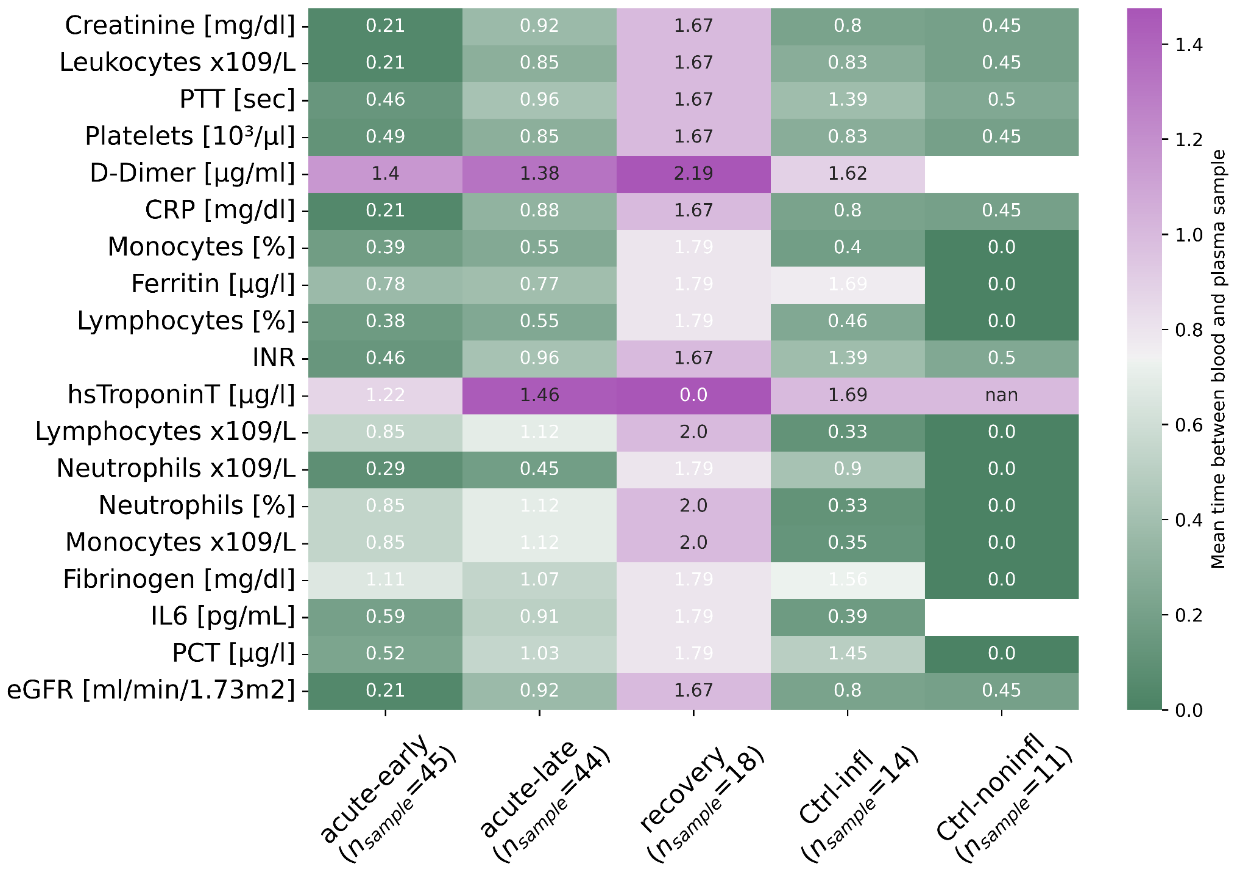


Color Code for the mean difference in days is shown on the right. The respective standard deviation for every laboratory value for the available information is shown as a number in every cell.

#### Figure S7: Missing laboratory values during trajectory

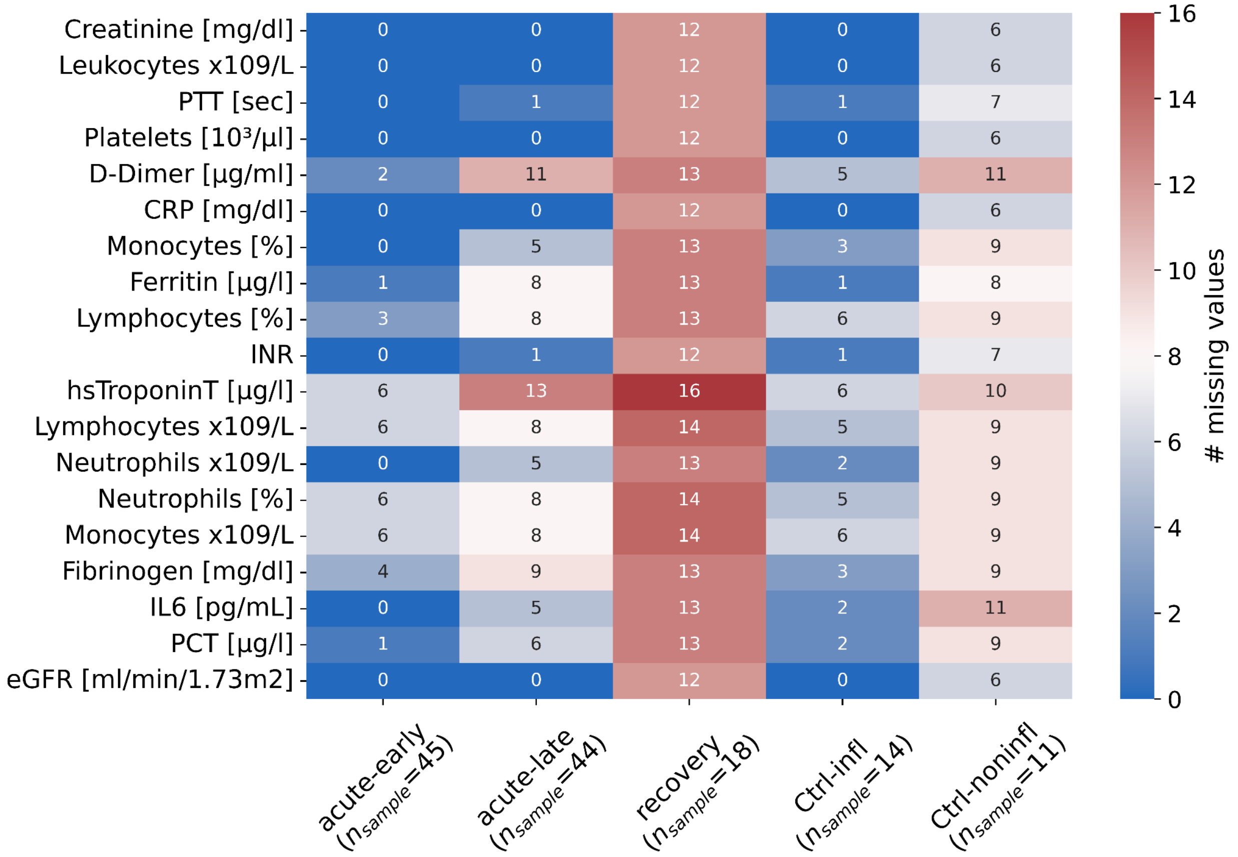
